# Predicting in-hospital indicators from wearable-derived signals for cardiovascular and respiratory disease monitoring: an in silico study

**DOI:** 10.1101/2025.03.11.25323722

**Authors:** Bianca Maria Laudenzi, Alberto Cucino, Sergio Lassola, Eleonora Balzani, Lucas Omar Müller

## Abstract

Cardiovascular and respiratory diseases (CVRD) are the leading causes of death worldwide. The construction of health digital twins for patient monitoring is becoming a fundamental tool to reduce invasive procedures, lower healthcare costs, minimize patient hospitalization, design clinical trials and personalize therapies. The aim of this study is to investigate the feasibility of machine learning-based monitoring of healthy subjects and CVRD patients in an in silico context. In particular, a population of virtual subjects, both healthy and with CVRD, was created using a comprehensive zero-dimensional global closed-loop model. Then, we trained Gaussian process regression (GPR) models, informed by wearable-acquired data, to predict variables normally acquired with invasive or operator-dependent methods. Presented results demonstrate, in an in silico setting, the feasibility of GRP-based prediction of in-hospital variables from wearable-derived indexes.

**Author summary:** Cardiovascular and respiratory diseases are major global health concerns. Effective remote monitoring is essential for early detection of complications and improved patient care, especially for people with chronic conditions. Wearable devices provide a non-invasive way to track health indicators, but they do not directly measure certain key physiological parameters that doctors typically assess in hospitals. In this study, we explore how machine learning approaches can help bridge this gap. By using virtually-generated data, we trained Gaussian process regression models to estimate critical cardiovascular and respiratory indexes, such as cardiac output and oxygen levels. In particular, we created a virtual population of simulated patients and used their data to train and test our model. Our findings suggest that this approach can be a valuable tool for remote monitoring, providing healthcare professionals with accurate insights, without the need for invasive procedures and enabling earlier detection of complications. However, further testing with real patient data is necessary to fully assess its clinical potential.

## Introduction

According to the World Health Organization [1], noncommunicable diseases account for more than 80 % of premature deaths. Cardiovascular and cardio-respiratory diseases (CVRD) account for the majority of non-communicable deaths (43 % and 10 % respectively), followed by cancer (21 %), and diabetes (4 %). Given the significant impact of CVRD on global mortality, effective monitoring of these diseases is essential for early detection and timely intervention. For this reason, the healthcare industry is witnessing an increase in the prevalence of wearable technologies since their impact on CVRD management has become undeniable [2–5]. The use of wearable technology in addressing CVRD offers significant clinical benefits such as decreasing healthcare costs, reducing the need for patients’ hospitalization, minimizing invasive procedures, designing clinical trials and personalized therapies, providing accurate long-term monitoring of cardiac indices during daily activities and sleep [3]. However, only a limited number of bio-signals can be estimated easily and accurately via commercial wearable devices, such as heart rate (HR), blood pressures (BPs), and arterial O_2_ saturation 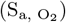 [4, 5], all derived using a photoplethysmogram (PPG), which is an optical non-invasive technique that uses a light-emitting diode to illuminate a capillary bed to monitor pulsatile changes in light absorption [6]. For many devices, their acquisition accuracy meets the FDA standards. Table 1 reports FDA-approved commercial wearable devices as well as their accuracy percentage for all wearable derived signals, i.e. HR, BPs, and 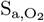. However, the clinical parameters required to monitor CVRD effectively extend beyond these signals. These include central venous pressure (CVP), stroke volume (SV), cardiac output (CO), left ventricular ejection fraction (EF), arterial partial pressure of 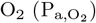, arterial partial pressure of 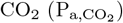.

**Table 1.**
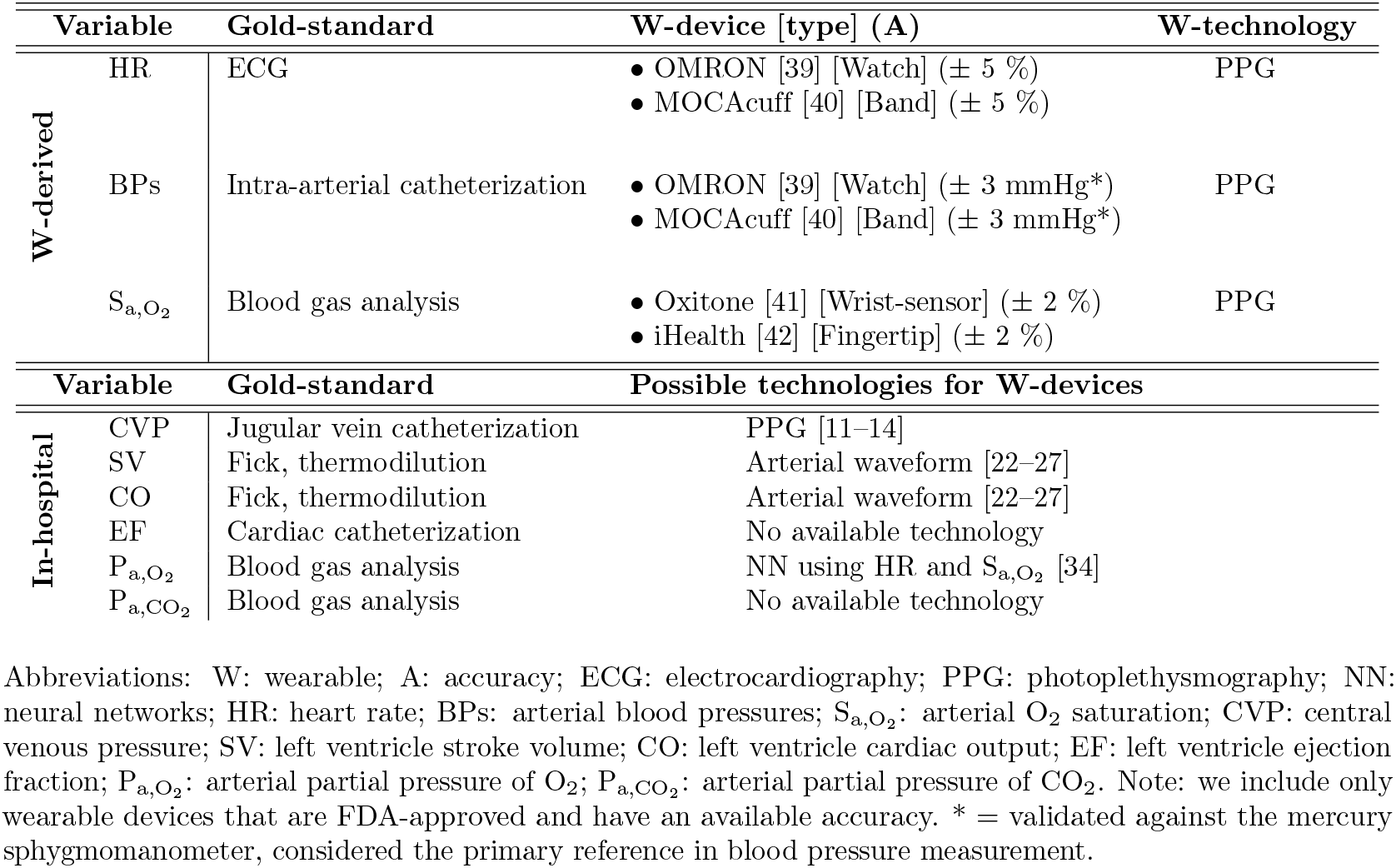
Summary of variables for CVRD monitoring and their acquisition methods.

CVP is used in clinical practice to assess volume status and cardiac preload, and its monitoring is crucial to understand and follow the hemodynamic status of patients with cardio-respiratory diseases. The gold standard technique for CVP measurement is invasive, requiring catheterization of the jugular vein [7]. However, well-accepted non-invasive options are ultrasound-guided techniques [8–10]. Some preliminary studies used PPG techniques to estimate CVP [11–14]. While promising, these findings were derived from relatively small sample sizes in specific surgical environments (perioperative or anesthesiology settings), indicating a need for broader validation across diverse clinical settings. Additionally, the need for precise positioning and multiple measurement points prevents existing methods from being automated and easily applicable, particularly in non-specialized home-based care, making this a significant limitation.

CO, SV, and EF are key indicators for evaluating cardiac circulatory failure and monitoring overall cardiac function [15, 16]. Methods for monitoring SV and CO are categorized into gold-standard invasive techniques (e.g., methods based on the Fick principle, thermodilution, pulse-indicated continuous CO method [17]) and non-invasive approaches (e.g., partial CO_2_ repeated respiration [18], nuclear magnetic resonance method [19], pulse contour analysis method [20] or studies on relationship between blood flow and skin temperature [21]). However, most of the latter are only suitable for single or short-term measurements and are not appropriate for 24-hour continuous and remote monitoring. More recent model-based methods [22–27] refine SV and CO estimation using features of the arterial peripheral waveform or more complex mathematical models based on waveform shape. Such approaches are promising for their implementation into wearable devices, but further development is needed to make these methods widely applicable in the wearable industry. EF can be measured invasively during cardiac catheterization by contrast left ventriculography [16], or non-invasively via imaging modalities such as echocardiography, TAC, or MRI [28–30]. Overall, the current non-invasive methods for EF remain operator-dependent and are unsuitable for use in a home setting.

Monitoring 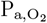 and 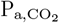 is essential to follow patients with acute respiratory failure such as hypercapnia and hypoxia, respectively [31]. Arterial blood gas analysis is the gold standard technique for such a purpose, but it is invasive, intermittent, and potentially painful [32]. Some studies estimate 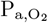 non-invasively and continuously using pulse rate and 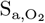, by means of nonlinear mixed effects regression analysis [33] or, more recently, neural networks [34]. However, both these studies collect data only from pediatric patients, limiting them to hospitalized conditions and small age ranges. Regarding 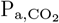, a non-invasive and remote technique uses transcutaneous sensors based on the principle of the Severinghaus electrode [35]. However, such monitors are not only expensive but also bulky and continuously drifting, requiring frequent recalibration by trained medical staff. Therefore, the current emphasis is on the development of wearable and easily deployable devices for remote patient monitoring outside clinical settings [32, 36].

From this literature review we have identified a set of clinically relevant variables for CVRD monitoring that are predominantly measured in hospital settings, each obtained through distinct acquisition techniques (i.e., CVP, CO, SV, EF, 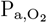, and 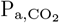). Throughout this text, we will refer to these as in-hospital variables. To the best of our knowledge, no comprehensive and general solution exists that predicts these in-hospital variables for remote monitoring. The challenge lies in predicting these clinical indicators remotely using a common methodology across all variables, leveraging wearable biosignals, such as HR, systolic and diastolic BPs, and 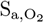, without requiring substantial new hardware or changing the acquisition technique of the wearable device. The development and validation of such methodology typically require a large set of measurements from real human subjects with sufficient variety. Such data collection can be a very arduous and expensive task. In this work we performed a preliminary assessment of the approach, without the need for extensive data collection, can be performed by utilizing virtual populations (VPs) that are generated through mathematical models, as demonstrated in similar studies [24, 37, 38].

## Materials and methods

The methodology followed in this work is summarized in Fig. 1. In particular, a virtual population was generated using a zero-dimensional (0D) global closed-loop cardio-respiratory model. This database of bio-signals enabled us to investigate the potential of Gaussian process regression (GPR) models in predicting in-hospital variables from wearable-acquired signals, in an in silico setting.

**Fig 1.**
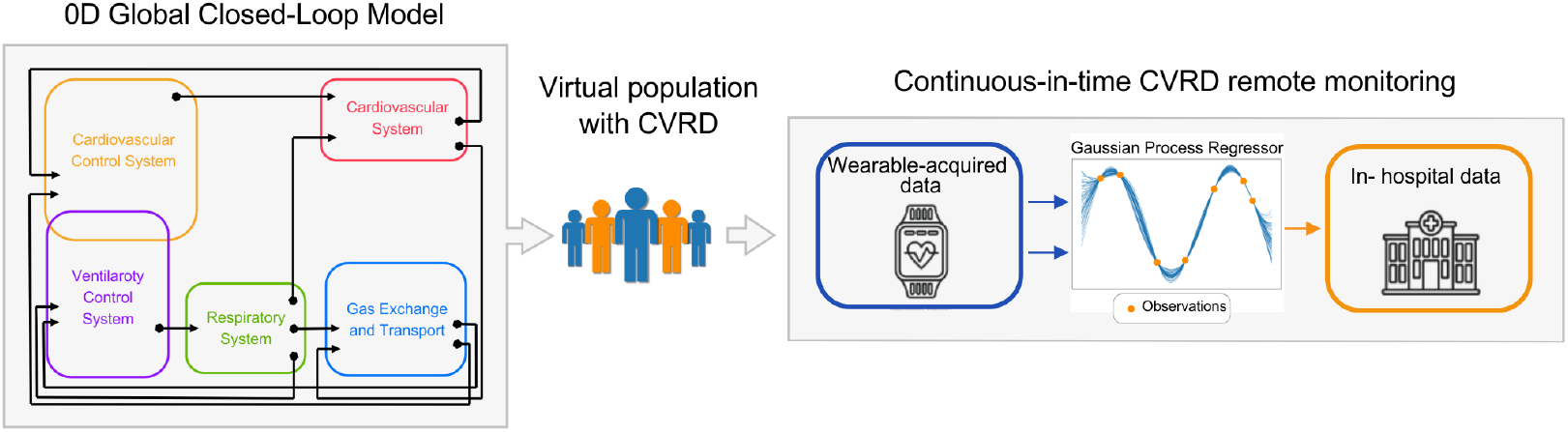
Graphical abstract. A virtual database is created using a comprehensive 0D global closed-loop cardio-respiratory model. Such database of bio-signals allowed us to explore Gaussian process regressor’s potential to predict in-hospital variables using wearable-acquired bio-signals.

### Global closed-loop cardio-respiratory model

The bio-signals of interest were simulated using a 0D global closed-loop mathematical model comprising major elements characterizing cardiovascular function, such as blood flow in heart chambers, systemic and pulmonary circulation, respiration and gas transport and metabolisms, as well as main short-term regulatory mechanisms. The selected model is an extension of model [43] and consists of a non-linear system of differential-algebraic equations. The model equations can be found in S1 Appendix. Fig. 2 shows a schematic representation of the cardio-respiratory model.

**Fig 2.**
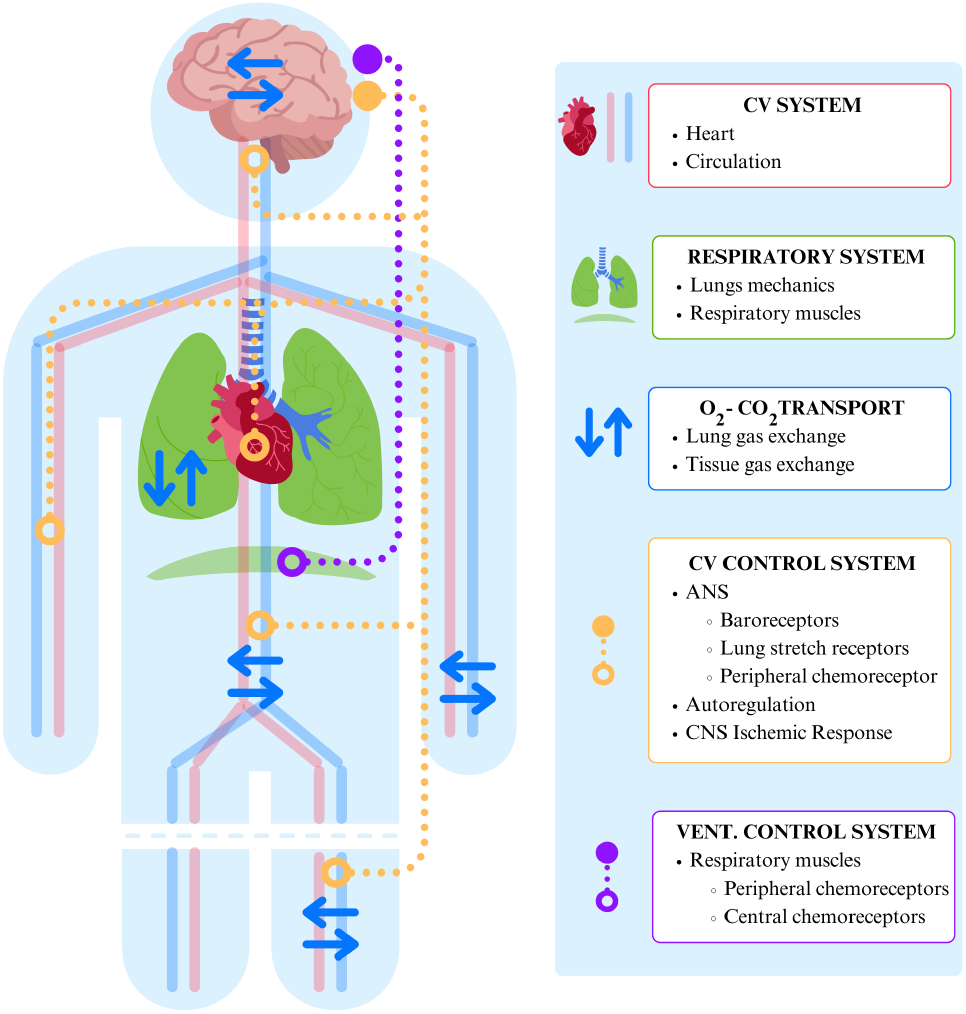
Schematic representation of the cardio-respiratory model. The figure shows the interaction between the cardiovascular and respiratory systems, highlighting gas exchange processes and control mechanisms. Abbreviations: CV: cardiovascular, ANS: autonomic nervous system, CNS: central nervous system, VENT.: ventilatory.

Numerical integration of differential equations was performed using the fourth-order Runge-Kutta method with a time step of 5 · 10^−5^ s, using an in-home code implemented in C++. For all simulations, model output signals were saved between 1940 and 2000 s, with a sampling period of 0.01 s. Clinically relevant variables were then extracted from this interval. Extracted model output variables included the wearable-derivable signals such as HR, central systolic/diastolic BPs (CSBP/CDBP), 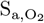, the in-hospital ones, i.e. CVP, SV, CO, EF, 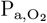, and 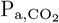 and other signals such as central pulse pressure (CPP), and mean arterial pressure (MAP). A precise description of the computation of the indexes is provided in S2 Appendix. Briefly, CDBP, MAP, and CSBP were identified respectively as the minimum, the mean, and the maximum of the arterial pressure waveform *P*_sa_ over each cardiac cycle. CPP was computed as the difference between CSBP and CDBP over each cardiac cycle. CVP, CO, 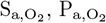 and 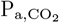 were computed respectively as the mean over each cardiac cycle of the thoracic vein pressure curve *P*_tv_, the flow curve through the aortic valve *q*_AV_, the arterial O_2_ saturation waveform 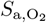, the arterial partial pressure curve of 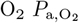, and the arterial partial pressure curve of 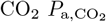. SV was computed as the product of CO and heart period for each cardiac cycle. Left ventricle end-diastolic and systolic volumes (LVEDV and LVESV) were identified respectively as the maximum and the minimum of the left ventricle volume curve *V*_LV_ over each cardiac cycle. EF was computed as the difference between LVEDV and LVESV over LVEDV, for each cardiac cycle. All indexes were averaged over the interval 1940–2000 s.

### Virtual database generation

A virtual database of clinically relevant variables for CVRD monitoring was generated by varying the most sensitive model input parameters. These parameters were identified through a local sensitivity analysis. Then we assigned uniform distributions to the selected parameters, using physiological and pathological ranges. The parameter space was then sampled effectively, and the cardio-respiratory model was run at each sampled point.

A post-processing filter was applied to ensure the physiological or pathological plausibility of the model output variables and that simulations reached a periodic state. In our simulations, cardiovascular and cardio-respiratory control mechanisms regulate key physiological variables, causing them to oscillate from one cardiac cycle to the next rather than following an identical periodic trajectory in every cardiac cycle. Therefore, we consider the system to be in a periodic state when the average values of clinically relevant variables, computed over a given time interval, remain stable over time.

CVRD-affected bio-signals were classified within the VP, ensuring the generation of both healthy and CVRD-affected patients.

#### Model parameters selection

For the identification of the most influential parameters, we explored the local sensitivity of clinically relevant model output variables to all 286 model input parameters. A simple definition of the local sensitivity 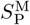 of model output M to changes in a certain parameter P is based on derivatives, i.e.

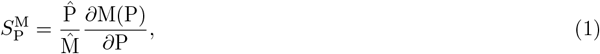

where 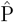 and 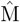 denote the baseline values for the considered parameter and model output [44]. In particular, we approximated the partial derivative in Eq. 1 using centered finite differences, and we chose to conduct the analysis by considering a 10 % variation below and above the baseline parameter value 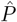.

To ensure a comprehensive representation of the cardio-respiratory system, we selected parameters influencing various physiological domains based on their significance in the local sensitivity analysis (see Table 6). The selected parameters are: the total blood volume (*V*_tot_) and the venous unstressed volume (*V*_u,ven_) since they ranked as the most relevant parameters for multiple clinically relevant outputs; the basal cardiac cycle (*T*_0_), the gain in vagal stimulus for heart period 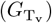, the onset of left atrium contraction 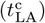 and the closing velocity of the mitral valve (*k*_close,MV_), since they affect EF; the carotid baroreflex activation level (*P*_n_) since it is the most influencial parameter representing a feedback mechanism; the inspired O_2_ fraction 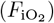, the hemoglobin content (*hgb*), the maximum O_2_ saturation (*C*_sat,O2_), and the empirical parameter for CO_2_ dissociation 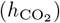, since they are the fundamental parameters describing gas exchange and transport. This selection ensured coverage of critical aspects of cardiovascular and respiratory physiology while focusing on parameters to which clinically relevant outputs are highly sensitive.

#### Generation of the virtual database

Parameters selected on the basis of local sensitivity analysis were considered as random variables with uniform distribution. The lower and upper bounds were based on ranges reported in the literature, as summarized in

Table 2. Specifically, the assignment of some ranges is based on the following assumptions. We assumed that the percentage variation of *V*_u,ven_ matched that of *V*_tot_. Given that the carotid baroreflex control system would typically adjust its set-point to maintain homeostasis in response to sustained changes in MAP, the baroreflex activation level *P*_n_ is assumed to vary proportionally with MAP. Furthermore, since the heart period increases linearly with efferent vagal frequency [45], it seemed physiologically reasonable to modify both the basal heart rate (*T*_0_) and the gain in vagal stimulus for the heart period 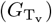 simultaneously.

**Table 2.** Physiological ranges of input parameters for the generation of VP 1.

| Parameter | Notation | Distribution | Units | Reference |
| --- | --- | --- | --- | --- |
| Total blood volume | $V_{tot}$ | $\mathcal{U}(4373.0, 6164.4)$ | mL | [46] |
| Venous unstressed volume | $V_{u,ven}$ | $\mathcal{U}(2574.9, 3629.6)$ | mL | [46] * |
| Basal cardiac cycle | $T_0$ *** | $\mathcal{U}(0.41, 0.59)$ | s | [47] |
| Gain in vagal stimulus for heart period | $G_{T_v}$ *** | $\mathcal{U}(0.07, 0.11)$ | $s \cdot \nu^{-1}$ | [45] |
| Baroreflex activation level | $P_n$ | $\mathcal{U}(81.9, 102.1)$ | mmHg | [47] ** |
| Onset of left atrium contraction | $t_{LA}^c$ | $\mathcal{U}(0.75, 0.85)$ | % of T | [48] |
| Closing velocity of mitral valve | $k_{close,MV}$ | $\mathcal{U}(37.3, 69.3)$ | $mmHg^{-1} \cdot s^{-1}$ | [49] |
| Inspired fraction of $O_2$ | $F_{iO_2}$ | $\mathcal{U}(14.7, 27.3)$ | % | [43] |
| Blood hemoglobin content | $hgb$ | $\mathcal{U}(0.15, 0.19)$ | $g \cdot mL^{-1}$ | [50] |
| Maximum concentration of hemoglobin-bound $O_2$ | $C_{sat,O_2}$ | $\mathcal{U}(1.7, 2.2)$ | $mL_{O_2} \cdot mL_{blood}^{-1}$ | [51] |
| Empirical parameters for $CO_2$ dissociation curve | $h_{CO_2}$ | $\mathcal{U}(1.7, 1.9)$ | - | [51] |

Note: $\nu$ = spikes/s; \* = we assumed that $V_{u,ven}$ has the same percentage variation as $V_{tot}$ ; \*\* = we assumed that the baroreflex activation level $P_n$ has the same variation of MAP; \*\*\* = we modify both $T_0$ and $G_{T_v}$ simultaneously [45].

We sampled the 10^th^-dimensional parameter space employing a quasi-Monte Carlo sampling strategy based on Sobol’ sequences with scrambling, implemented via the *qmc*.*Sobol* function from the *SciPy* library. This method generates low-discrepancy samples in the multidimensional parameter space, ensuring uniform coverage of the domain [52]. The cardio-respiratory model was executed for every point in the sampled parameter space. Convergence of the number of sampling points used to generate the VP was verified running the model with 128, 1024, 4096, 8192 sampling points and comparing means, standard deviations (SDs), minimums and maximums of all clinically relevant variables. The errors for convergence were computed with respect to the highest number of sampling points as

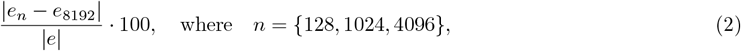

and *e* represents either the mean, the SD, the minimum, or the maximum of a specific variable.

#### Filter criteria and CVRD classification

Following [53], we applied a post-processing filter to ensure that each numerical simulation reached a periodic state and remained within the physiological ranges representative of healthy or CVRD-affected individuals. In particular, we excluded simulations meeting either one of the following criteria: (1) any clinically relevant model output variable showed a difference greater than 1 % between the data collected in the intervals 1940–2000 s and 2940–3000 s of the simulation, or (2) any variable fell outside its specified physiological or pathological range. The admissible ranges for each variable are provided in Table 3.

**Table 3.** Filter criteria for model output variables.

| Variable | Min | Max | Unit | Reference |
| --- | --- | --- | --- | --- |
| HR | 45 | 100 | beats · min<sup>-1</sup> | [47] |
| CSBP | 84 | 174 | mmHg | [47] |
| CDBP | 59 | 110 | mmHg | [47] |
| CPP | 13 | 86 | mmHg | [47] |
| MAP | 74 | 132 | mmHg | [47] |
| CVP | 0 | 10 | mmHg | [47] |
| SV | 44 | 128 | mL | [54] |
| CO | 42 | 150 | mL · s<sup>-1</sup> | [54] |
| EF | 45 | 74 | % | [55] |
| S<sub>a,O<sub>2</sub></sub> | 80 | 100 | % | [56] |
| P<sub>a,O<sub>2</sub></sub> | 70 | 110 | mmHg | [56] |
| P<sub>a,CO<sub>2</sub></sub> | 30 | 50 | mmHg | [56] |

Abbreviations: HR: heart rate; CSBP/CDBP: central systolic/diastolic blood pressure; CPP: central pulse pressure; MAP: mean arterial pressure; CVP: central venous pressure; SV: left ventricle stroke volume; CO: left ventricle cardiac output; EF: left ventricle ejection fraction; S<sub>a,O<sub>2</sub></sub>: arterial O<sub>2</sub> saturation; P<sub>a,O<sub>2</sub></sub>: arterial partial pressure of O<sub>2</sub>; P<sub>a,CO<sub>2</sub></sub>: arterial partial pressure of CO<sub>2</sub>.

Furthermore, since we wanted to create a virtual database representative of both healthy and CVRD-affected patients, we classified simulations that passed the filtering stage into healthy or CVRD-affected bio-signals, such as bradycardia, hypertension, hypo and hyperventilation and reduced cardiac function, according to the cutoff values reported in Table 4. Notably, a single simulation could be classified under multiple pathological conditions.

**Table 4.** CVRD-classification for model output variables.

| Variable | Cutoff | Unit | CVRD | Reference |
| --- | --- | --- | --- | --- |
| HR | < 60 | beats · min<sup>-1</sup> | Bradycardia | [57] |
| CSBP | > 130 | mmHg | Hypertension | [58] |
| CDBP | > 85 | mmHg | Hypertension | [58] |
| EF | < 50 | % | Reduced cardiac function | [59] |
| P<sub>a,CO<sub>2</sub></sub> | < 35 | mmHg | Hyperventilation | [58] |
| P<sub>a,CO<sub>2</sub></sub> | > 45 | mmHg | Hypoventilation | [58] |

Abbreviations: HR: heart rate; CSBP/CDBP: central systolic/diastolic blood pressure; EF: left ventricle ejection fraction; P<sub>a,CO<sub>2</sub></sub>: arterial partial pressure of CO<sub>2</sub>.

### Prediction of in-hospital variables

#### Gaussian process regression

We used GPR for predicting in-hospital data by leveraging bio-signals that can be acquired through wearable devices, in accordance with the literature review summarized in Table 1. Briefly, a GPR model is a statistical model that describes how a scalar simulator output *f* (**x**) varies as a function of *D* input features **x** = (*x*_1_, …, *x*_*D*_). The simulator outputs, *f* (·), are modeled as being jointly Gaussian with prior mean

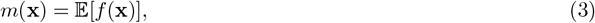

and covariance

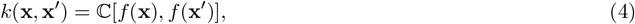

where *k*(**x, x**^*′*^) is a positive-definite kernel [60]. In this case, the GPR model training is done in Python with *Scikit Learn* [61]. For each GRP model, the kernel is the product of a Constant kernel (*sklearn*.*gaussian process* .*kernels*.*ConstantKernel*) and a Radial Basis Function kernel (*sklearn*.*gaussian process*.*kernels*.*RBF*), in which the hyperparameters are tuned using the function *sklearn*.*model selection*.*RandomizedSearchCV* with 10-fold cross-validation with random shuffling of the data and maximum error scoring strategy (*sklearn*.*metrics*.*max error*). After selecting the simulations that have not failed the filter criteria, the virtual dataset VP 1 is divided into 80 % training (1332 samples) and 20 % test (333 samples). We train independently six GPR models on the normalized training set 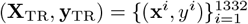, where **X**_TR_ is the input features matrix of wearable-derived signals, with

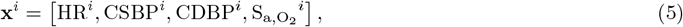

while **y**_TR_ represents the vector of one of the six target variables that we aim to predict independently, i.e.

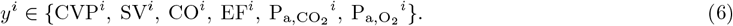

The performance of the GPR models were then evaluated on the test set 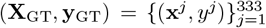, exploiting the standard scoring metrics: the coefficient of determination *R*^2^, the percentage maximum relative error, i.e.

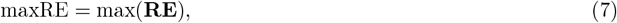

and the percentage mean relative error, i.e.

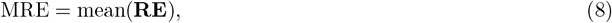

where **RE** is the percentage relative error.

To assess the effect of the training set size on the performance of the GPR models, learning curves were computed for training sizes of 100, 200, 300, 500, and 1000 data points, sampled from the VP 1 train set. For this purpose, we used the *sklearn*.*model selection*.*learning curve* function with 10-fold cross-validation with random shuffling of the data and maximum error scoring strategy (*sklearn*.*metrics*.*max error*).

#### Error propagation analysis

The input features provided to the GPR models are signals acquired through wearable devices, which inherently include acquisition errors. In order to account for this in our work, we assumed acquisitions errors of *±* 5 % for HR [39, 40] and ± 2 % for 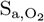 [41, 42]. For BP signals we needed to account not only for the errors introduced by the wearable devices but also for the errors resulting from estimating central BP from peripheral BP measurements using transfer functions. Indeed, while the cardio-respiratory model accurately represents central BPs, which are the ones used in the input vector features, i.e. CSBP and CDBP, it does not provide precise peripheral BP values. The mean bias of transfer functions, compared to gold-standard measurements, ranges between 1-5 mmHg [62–64], whereas the error introduced by the wearable-device acquisition process is ± 3 mmHg [39, 40]. As a result, we assumed an overall error of ± 3 mmHg for CSBP and CDBP. Given these acquisition errors, we conducted an error propagation analysis for each trained GPR model to assess the impact of these errors on the model’s performance.

In particular, assuming that the errors are distributed uniformly, for each test data point *j*, with *j* = 1, …, 333, we sampled *N* -times the multidimensional uniform distribution of input features with their acquisition errors, using a quasi-Monte Carlo sampling strategy based on Sobol’ sequences with scrambling, implemented via the *qmc*.*Sobol* function from the *SciPy* library. Thus, we obtained the sample set of vectors **𝒳**^*j*^, i.e.

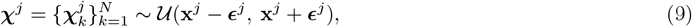

where

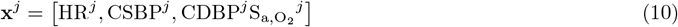

is the input features vector for the test data point *j* and

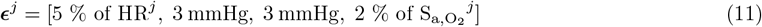

is the corresponding input reference acquisition errors ranges.

Finally, for each test data point *j*, we evaluated the six trained GPR models on the sample set **𝒳**^*j*^, obtaining the corresponding sample set of predictions 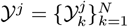, with

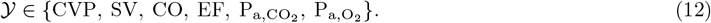

The performance of the GPR models predictions with perturbed input features was evaluated on the test set 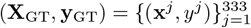 using the following metrics. The percentage maximum relative error

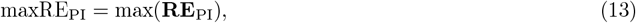

the percentage mean relative error

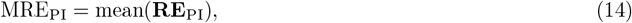

and the coefficient of variation

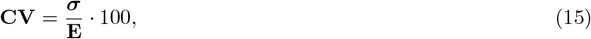

where **RE**_PI_ is the vector of percentage relative errors with perturbed input features defined as

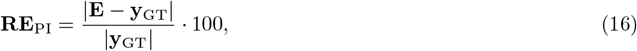

with 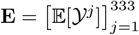 being the vector of estimated expected values of 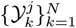 and 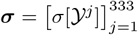 representing the vector of estimated SDs of 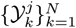.

Different sampling sizes *N* = {50, 100, 500, 1000} of 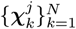 were used to ensure convergence of the methodology comparing means and SDs of the vector of percentage relative errors **RE**_PI_.

#### Validation against new virtual population

To ensure that the predictive capabilities of our model were not overly reliant on the parameters choice that we made for the generation of VP 1, we tested the GPR models, which were trained on the VP 1 dataset, on a new dataset. This new population was called VP 2 and was obtained by varying different combinations of 0D model parameters. In this case, we selected the model input parameters analyzing again the results of the local sensitivity analysis (see Table 6) but using a different criteria: we chose all the parameters that ranked in the first 3 ranking positions, i.e. the total blood volume (*V*_tot_), the venous unstressed volume (*V*_u,ven_), the basal cardiac cycle (*T*_0_), the gain in vagal stimulus for heart period 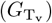, the onset of left atrium contraction 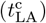, the inspired fraction of 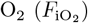, the blood hemoglobin content (*hgb*), the left and right passive ventricles elastances (*k*_E,LV_, *k*_E,RV_), the maximum concentration of hemoglobin-bound 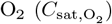 and the empirical parameters for the O_2_/CO_2_ dissociation curve 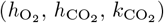. Following the same procedure as before, we assigned uniform distributions to the selected model parameters, based on the physiological and pathological ranges summarized in Table 5, then we sampled the parameter space employing the *qmc*.*Sobol* function from the *SciPy* library. The cardio-respiratory model was run at each sampled point. Note that the basal heart rate *T*_0_ and the gain in vagal stimulus for the heart period were modified simultaneously, as well as the left and right passive ventricles elastances (*k*_E,LV_ and *k*_E,RV_), resulting in a 11^*th*^-dimensional parameter space.

**Table 5.** Physiological ranges of input parameters for the generation of VP 2.

| Parameter | Notation | Distribution | Unit | Reference |
| --- | --- | --- | --- | --- |
| Total blood volume | $V_{\text{tot}}$ | $\mathcal{U}(4373.0, 6164.4)$ | mL | [46] |
| Venous unstressed volume | $V_{\text{u,ven}}$ | $\mathcal{U}(2574.9, 3629.6)$ | mL | [46] * |
| Basal cardiac cycle | $T_0$ ** | $\mathcal{U}(0.41, 0.59)$ | s | [47] |
| Gain in vagal stimulus for heart period | $G_{T_v}$ ** | $\mathcal{U}(0.07, 0.11)$ | $\text{s} \cdot \nu^{-1}$ | [45] |
| Onset of left atrium contraction | $t_{\text{LA}}^c$ | $\mathcal{U}(0.75, 0.85)$ | % of T | [48] |
| Inspired fraction of $\text{O}_2$ | $F_{\text{iO}_2}$ | $\mathcal{U}(14.7, 27.3)$ | % | [43] |
| Blood hemoglobin content | $hgb$ | $\mathcal{U}(0.15, 0.19)$ | $\text{g} \cdot \text{mL}^{-1}$ | [50] |
| Passive elastance of left ventricle | $k_{\text{E,LV}}$ *** | $\mathcal{U}(0.01, 0.02)$ | $\text{mL}^{-1}$ | [65] |
| Passive elastance of right ventricle | $k_{\text{E,RV}}$ *** | $\mathcal{U}(0.008, 0.014)$ | $\text{mL}^{-1}$ | [65] |
| Maximum concentration of hemoglobin-bound $\text{O}_2$ | $C_{\text{sat,O}_2}$ | $\mathcal{U}(1.7, 2.2)$ | $\text{mL}_{\text{O}_2} \cdot \text{mL}_{\text{blood}}^{-1}$ | [51] |
| Empirical parameter for $\text{O}_2$ dissociation curve | $h_{\text{O}_2}$ | $\mathcal{U}(0.38, 0.39)$ | - | [51] |
| Empirical parameter for $\text{CO}_2$ dissociation curve | $h_{\text{CO}_2}$ | $\mathcal{U}(1.7, 1.9)$ | - | [51] |
| Empirical parameter for $\text{CO}_2$ dissociation curve | $k_{\text{CO}_2}$ | $\mathcal{U}(110, 279)$ | - | [51] |

Note: $\nu$ = spikes/s; \* = we assumed that $V_{\text{u,ven}}$ has the same percentage variation as $V_{\text{tot}}$ ; \*\* = $T_0$ and $G_{T_v}$ are modified simultaneously; \*\*\* = $k_{\text{E,LV}}$ and $k_{\text{E,RV}}$ are modified simultaneously.

The post-processing filter criteria defined in Table 3 as well as the CVRD classification scheme reported in Table 4 were applied. After selecting the simulations that did not fail the filter criteria, the six GPR models were tested on VP 2. Following the same procedure described in Section Error propagation analysis, we tested the GPR models with perturbed input data of VP 2.

## Results

First, we examine output variables from the 0D cardio-respiratory model, focusing on validation, feature extraction, and local sensitivity analysis. Then, we address the results of the VPs, focusing on convergence, global sensitivity analysis, variables distributions and post-processing filters, as well as a comparison between VP 1 and VP 2. Lastly, we explore performance of the GPR models predictions for the in-hospital variables.

### Cardio-respiratory model

The 0D model was validated in the baseline case by comparing the model-predicted cardiac and hemodynamic indices with reference values reported in the literature. For a detailed comparison between the model predictions and the literature data, please refer to S1 Appendix.

Fig. 3 shows model output variables over time (blue lines) and extracted features (orange indexes) for the 0D cardio-respiratory model in the baseline case. The first plot displays the heart cycle duration, *T*, which exhibits the expected variations due to the interaction of sympathetic and vagal activity over the shorter timescale and respiratory modulation over the longer timescale. The range values close to 1 s are consistent with a normal resting heart rate [47]. The dimensionless variable *u* represents the fraction of the cardiac cycle and allows for the identification of each heart period, with *u* = 0 assumed at the beginning of systole. The systemic arterial pressure, *P*_sa_, displays a physiological pulsatile pattern, with extracted features for CSBP, CDBP, and MAP all within normal ranges [47]. Similarly, the thoracic vein pressure, *P*_tv_, reflects the expected CVP range with small oscillations around a stable baseline value [66]. The flow through the aortic valve, *q*_AV_, shows a typical systolic ejection phase, with a slightly elevated peak flow, along with a brief period of backward flow at the beginning of diastole. This behavior is due to including non-ideal valves in the model, representing a modification compared to the previous approach [43] that assumed ideal valves. The left ventricular volume, *V*_LV_, follows the expected pattern of filling and emptying of the ventricle, with extracted features for LVEDV and LVESV volumes yielding a normal stroke volume [67]. The arterial oxygen saturation, 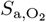, shows small oscillations around a stable baseline value, while the partial pressures of oxygen 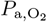 and carbon dioxide 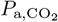 oscillate within physiological ranges due to respiratory cycles [56].

**Fig 3.**
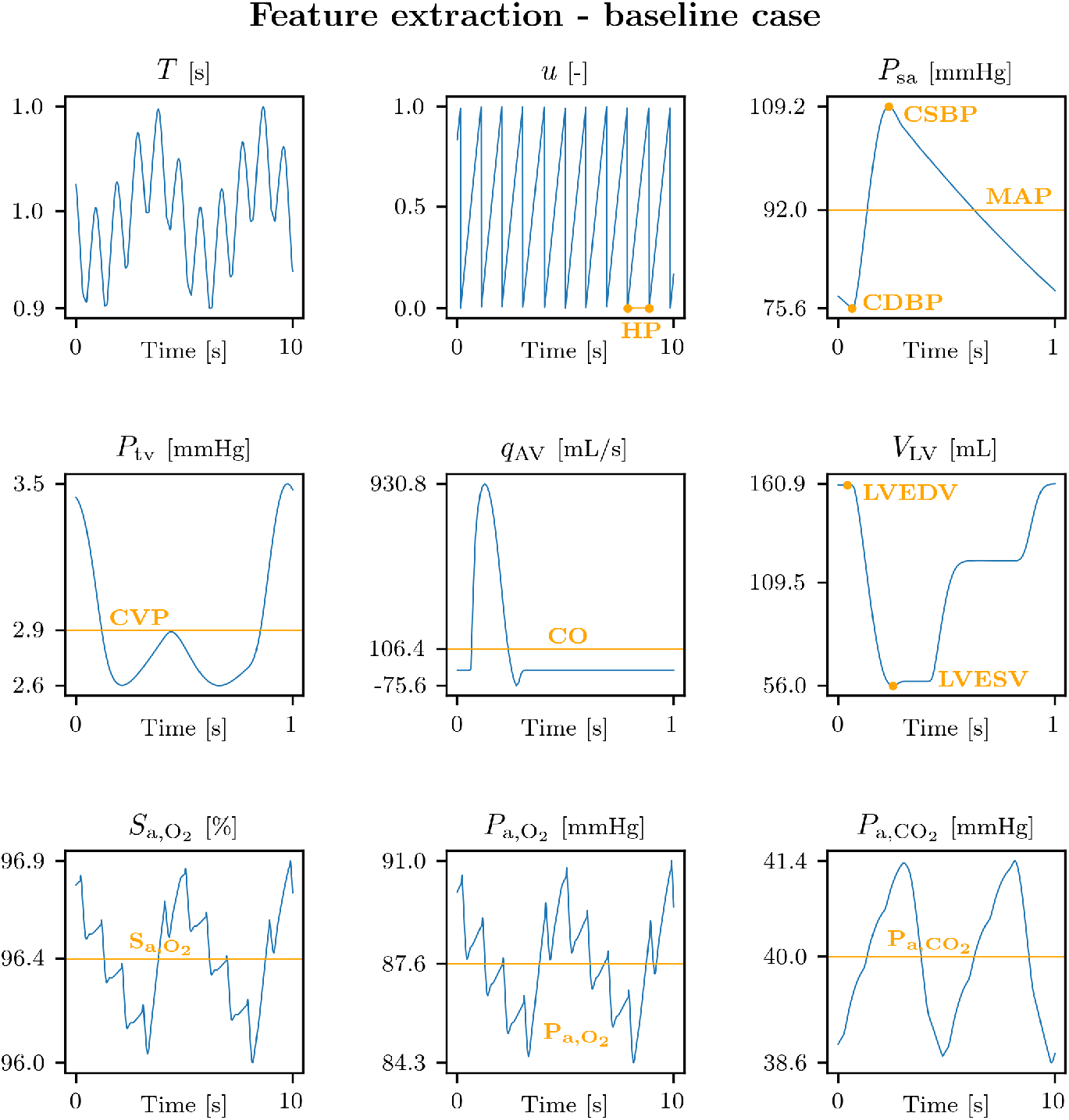
Cardio-respiratory model features extraction. Signals over time (blue curves) and extracted features (orange indexes). The y-axes indicate the minimum, the mean, and the maximum values for each signal. Abbreviations: *T* : heart cycle duration; *u*: fraction of the cardiac cycle; *P*_sa_: systemic arterial pressure; *P*_tv_: thoracic vein pressure; *q*_AV_: flow though the aortic valve; *V*_LV_: left ventricle volume; 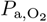: arterial O_2_ saturation; 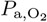 arterial partial pressure curve of O_2_; 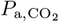: arterial partial pressure curve of CO_2_; HP: heart period; CSBP/CDBP/MAP: central systolic/diastolic/mean blood pressure; CVP: central venous pressure; CO: left ventricle cardiac output; LVESV/LVEDV: left ventricle end systolic/diastolic volume; 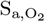: arterial O_2_ saturation; 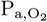: arterial partial pressure of O_2_; 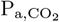: arterial partial pressure of CO_2_.

Table 6 shows the local sensitivity analysis results of all 286 model inputs for some selected model outputs, i.e. the wearable-acquirable signals HR, CSBP, CDBP, and 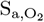, and the in-hospital ones, CVP, SV, CO, EF, 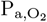, and 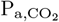. Only the parameters ranked within the first five ranking positions, together with their sensitivity index 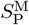, are reported. The total blood volume *V*_tot_ and the venous unstressed volume *V*_u,ven,_ appear to be the most influential parameters across all the cardiovascular signals, such as HR, CSBP, CDBP, CVP, SV and CO, with the exception of EF, for which the time constants 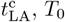 and 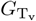 appear in the first positions of the rankings. While the cardiovascular variables are predominantly affected by the same parameters, each cardio-respiratory variable, i.e. 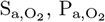 and 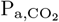, is primarily sensitive to a distinct parameter. Among these, 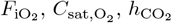 and *hgb* stand out as the most prominent parameters in determining their behavior. These findings inform the selection of key parameters for generating VP 1 and VP 2.

### Virtual populations

In this section, we report the results of VP 1, similar results for VP 2 are omitted here for brevity but can be found in S3 Appendix.

Fig. 4 illustrates the convergence of the variables of interest for different sampling point numbers (128, 1024, 4096, and 8192), specifically reporting percentage errors of means, SDs, minimums, and maximums for all clinically relevant variables of VP 1. For the means and SDs, strong convergence was observed across all variables, with maximum error variations of 0.01 % and 0.1 %, respectively, at 4096 sampling points. However, convergence was more challenging to achieve for minimums and maximums. In particular, the minimums showed a maximum error across all variables of 7 % for 4096 sampling points. In contrast, the maximums exhibited better convergence, with a maximum error of 1 %. Overall, we conclude that 4096 sampling points are sufficient to ensure convergence, as the errors for all statistical indicators remain acceptably low.

**Fig 4.**
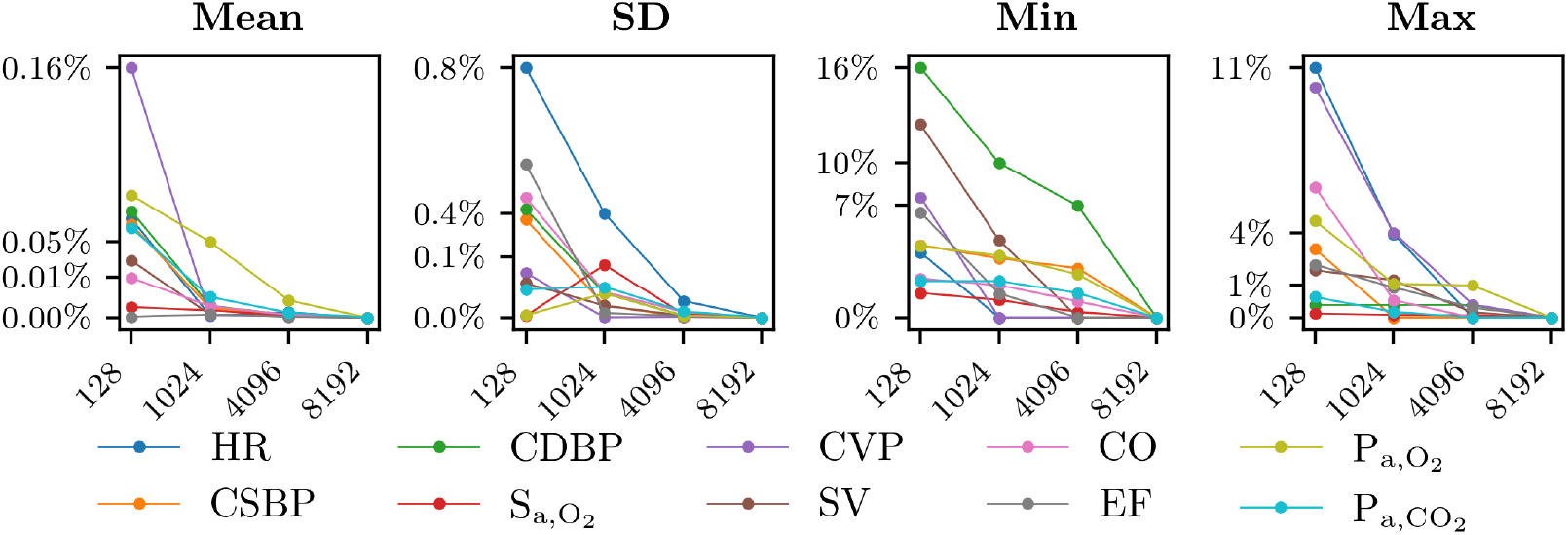
Convergence of the number of sampling points. Percentage errors of means, standard deviations (SDs), minimums and maximums for all variables of interest of VP 1 (y-axes), for different sampling points sizes (x-axes). Abbreviations: HR: heart rate; CSBP/CDBP: central systolic/diastolic blood pressure; 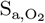: arterial O_2_ saturation; SV: left ventricle stroke volume; CO: left ventricle cardiac output; EF: left ventricle ejection fraction; 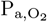: arterial partial pressure of O_2_; 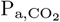: arterial partial pressure of CO_2_.

A global sensitivity analysis was performed on the 10 parameters varied for generating VP 1, using the *analyze*.*enhanced hdmr*.*analyze* function form the *SALib* library [68]. Fig. 5 shows the heat map of the total Sobol indices for the selected 10 model input parameters over all clinically relevant output variables of VP 1 generated with 4096 sampling points. Darker colors refer to higher interaction between inputs and outputs. The indexes for each parameter were normalized so that they sum up to 1, representing 100 % of the output variance. Indexes lower than 0.01 were not annotated in the heat map. The variance-based global sensitivity analysis identified the total blood volume, *V*_tot_, and the venous unstressed volume, *V*_u,ven_, as the most significant parameters for a subset of selected signals of the cardiovascular system, i.e. HR, CSBP, CDBP, CVP, SV, CO, EF. Moreover, the basal heart period, *T*_0_, together with the gain in vagal stimulus for heart period, 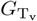, also had a significant influence on HR and EF. On the other hand, the inspired fraction of 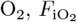, and the O_2_ saturation concentration, 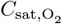, were the most significant parameters for the cardio-respiratory bio-signals, i.e. 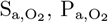 and 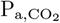. These results confirm the relevance of some parameters identified in the local sensitivity analysis, while others occupy different ranks. In particular, parameters such as 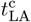 and 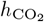, which ranked high in local sensitivity, appeared less influential in global sensitivity. This difference can be explained by the smaller variation (±6 % from the baseline value) used for generating VP 1, compared to the broader variation (±10 % from the baseline) applied in the local sensitivity analysis, as well as by the role of parameters interactions within the model, which are not captured in the local sensitivity analysis.

**Fig 5.**
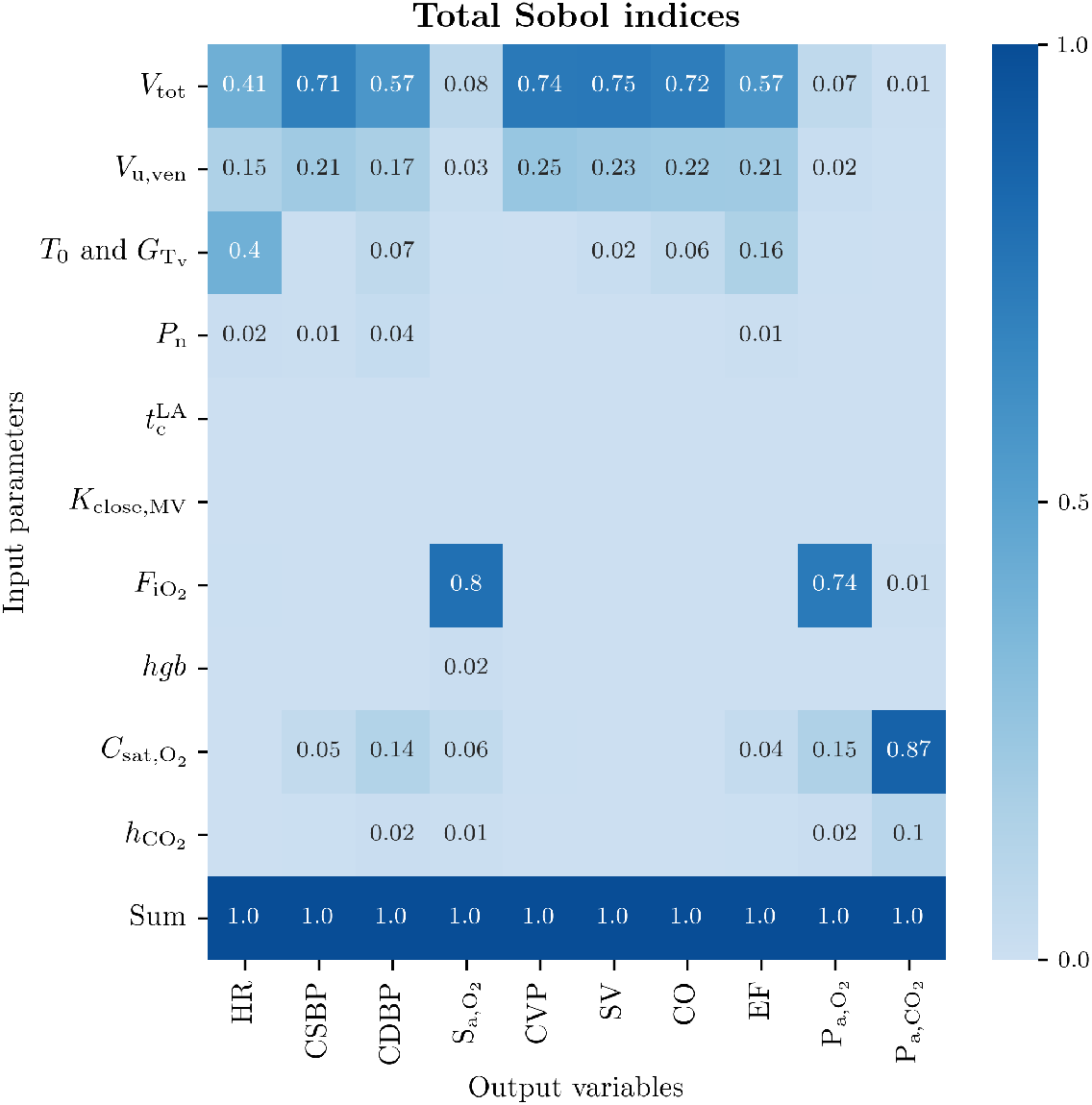
Global sensitivity analysis. Total effects of input parameters (y-axis) on model outputs (x-axis), normalized between 0 and 1, for VP 1. Parameters abbreviations: *V*_tot_: total blood volume; *V*_u,ven_ venous unstressed volume; *T*_0_: basal cardiac cycle; 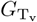: gain in vagal stimulus for heart period; *P*_n_: baroreflex activation level; 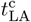: onset of left atrium contraction; *k*_close,MV_: closing velocity of mitral valve; 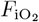: inspired fraction of O_2_; *hgb*: blood hemoglobin content; 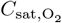: maximum concentration of hemoglobin-bound O_2_; sh: pulmonary shunt; 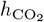: empirical parameters for CO_2_ dissociation curve. Variables abbreviations: HR: heart rate; CSBP/CDBP: central systolic/diastolic blood pressure; 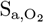: arterial O_2_ saturation; SV: left ventricle stroke volume; CO: left ventricle cardiac output; EF: left ventricle ejection fraction; 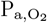: arterial partial pressure of O_2_; 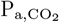: arterial partial pressure of CO_2_.

Fig. 6 shows the distributions of the model outputs of VP 1 generated with 4096 sampling points. The translucent blue bands over the histograms indicate that the value of that specific variable did not meet the filter criteria detailed in Table 3. The simulations with output variables falling within those regions were excluded from the VP. The other translucent colored bands (red, orange, green, yellow, and purple) refer to the different CVRD according to the cutoff values reported in Table 4. The shapes of the distributions vary depending on the physiological variable, with some showing clear peaks (e.g., HR, EF), while others exhibit more uniform or flat patterns (e.g., CPP, SV, CO). All variables are well distributed within their respective reference ranges, with the exception of 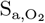, where a broader range would have been preferable. Indeed, the filter criteria are not applied to all variables of VP 1, particularly for EF and 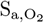, where the cutoff values in Table 3 are never met. Similarly, upper cutoff values for CPP, MAP, CVP, and 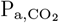 were never reached in pre-filtered VP1 subjects. In particular, Fig. 7 shows, on the left, a pie chart illustrating which simulations failed the post-processing filter critera out of all 4096 simulations. On the right, a second pie chart shows the percentage of simulations that failed the filter criterion for each output variable, among the simulations that failed the filter criteria. Out of 4096 simulations, 2431 failed the filter criteria, therefore the final VP 1 is composed of 1665 subjects. Additionally, we note that the majority of failed simulations are due to too high 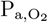, too low MAP, CSBP and CDBP, or too low CVP. Furthermore, it has been verified that a large portion of rejected samples is due to incompatible combinations of input parameters. For instance, a low value of *V*_tot_ combined with a high value of *V*_u,ven_ leads to excessively low MAP, while the opposite case, a high value of *V*_tot_ combined with a low value of *V*_u,ven_, results in an excessively high SV. population.

**Fig 6.**
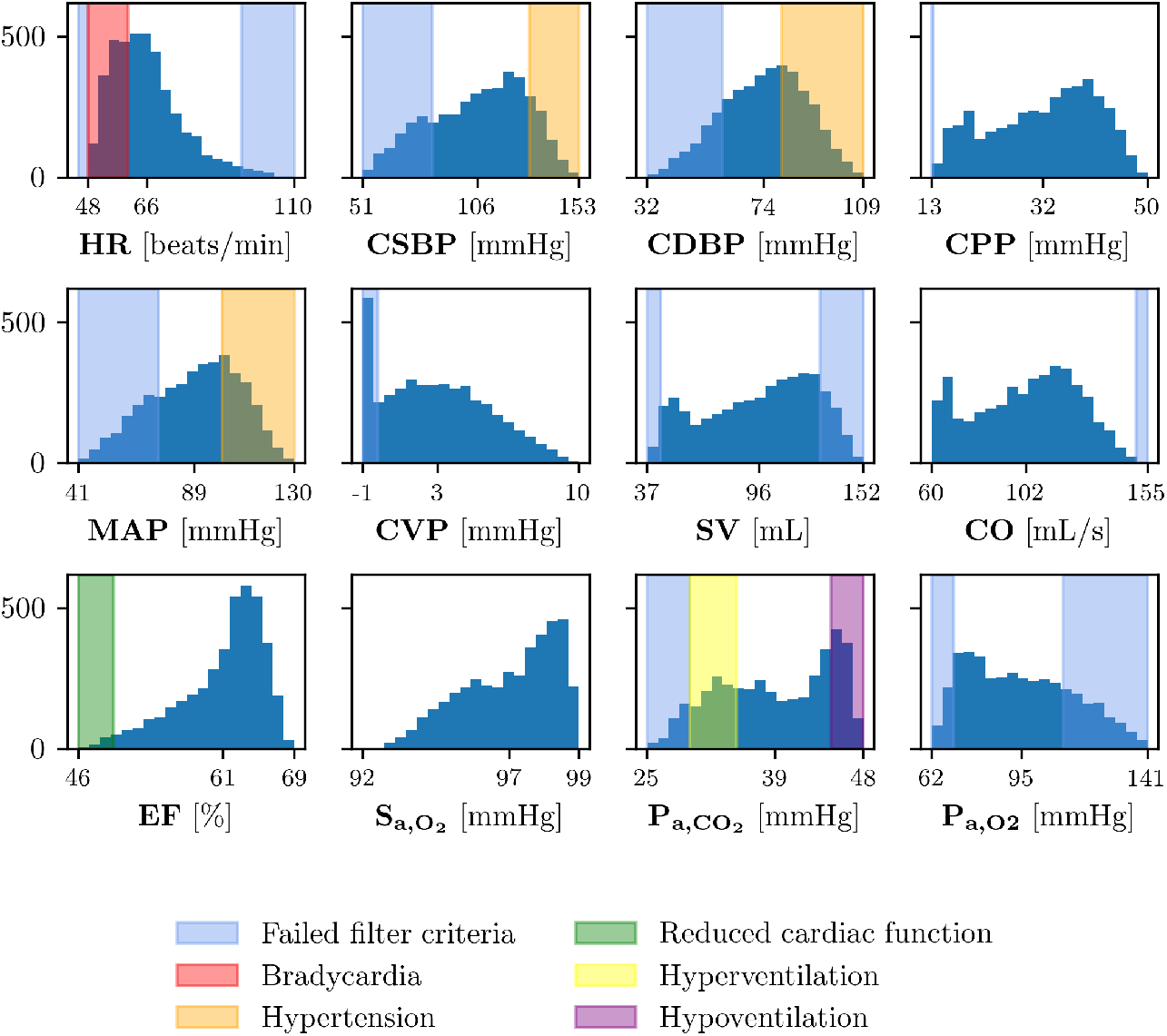
Virtual population distributions. Frequency (y-axes) of values of all model outputs (x-axes) for VP 1. The minimum, the mean, and the maximum of each variable are shown in the x-axes. Translucent blue bands indicate that the variable does not meet the filter criteria; translucent colored bands (red, orange, green, yellow, and purple) refer to the different CVRD. Abbreviations: HR: heart rate; CSBP/CDBP: central systolic/diastolic blood pressure; CPP: central pulse pressure; MAP: mean arterial pressure; CVP: central venous pressure; SV: left ventricle stroke volume; CO: left ventricle cardiac output; EF: left ventricle ejection fraction; 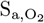: arterial O_2_ saturation; 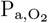: arterial partial pressure of O_2_; 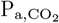: arterial partial pressure of CO_2_.

**Fig 7.**
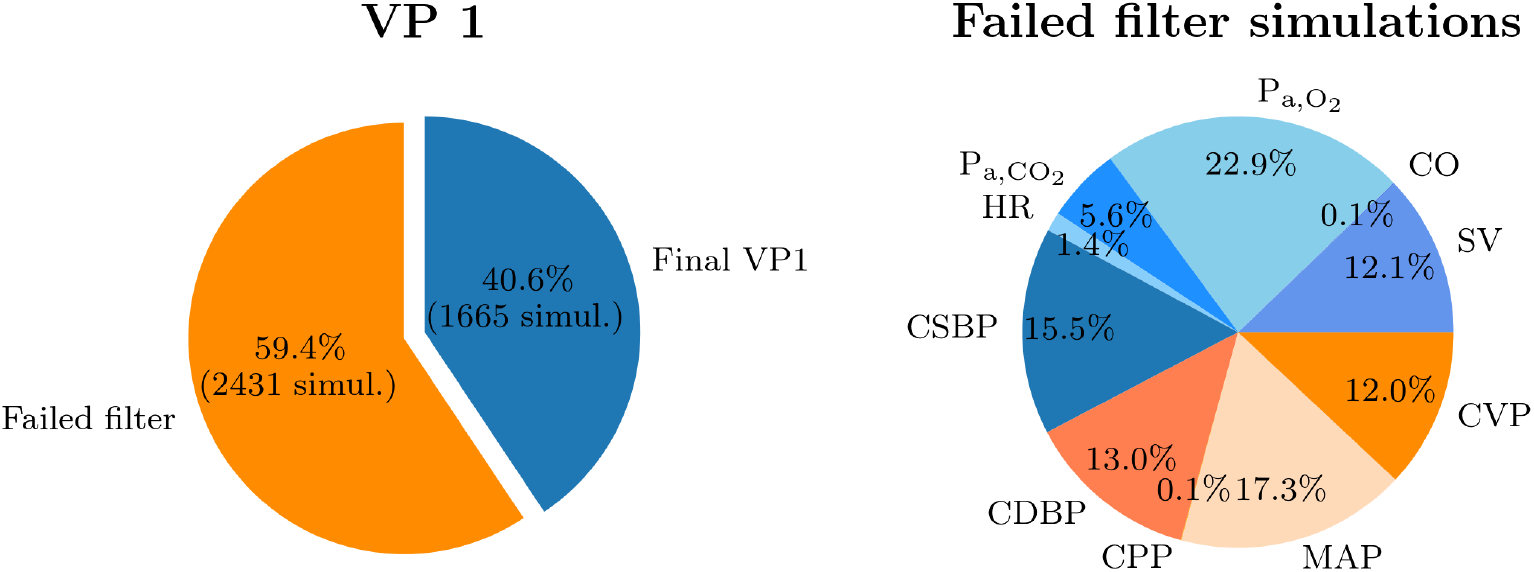
Post-processing filter criteria. On the left, the simulations that have failed the post-processing filter criteria among all 4096 simulations of VP 1. On the right, the percentage of failed simulations, for each output variable, among the simulations that have failed the filter criteria of VP 1. Abbreviations: HR: heart rate; CSBP/CDBP: central systolic/diastolic blood pressure; CPP: central pulse pressure; MAP: mean arterial pressure; CVP: central venous pressure; SV: left ventricle stroke volume; CO: left ventricle cardiac output; EF: left ventricle ejection fraction; 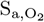: arterial O_2_ saturation; 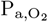: arterial partial pressure of O_2_; 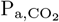: arterial partial pressure of CO_2_.

Fig. 8 shows the pie-chart of the CVRD classification among the simulations that have not failed the filter criteria. Among those, we can distinguish between healthy subjects (33.8 %), hypertensive patients (24.0 %), hyper- or hypo-ventilating ones (17.5 % and 6.3 % respectively), or combinations of those. We note that, although all CVRD were initially present before applying the filter criteria (Fig. 6), not all of them remain in the final VP. For instance, bradycardia and reduced ejection fraction are excluded after the filter criteria are applied.

**Fig 8.**
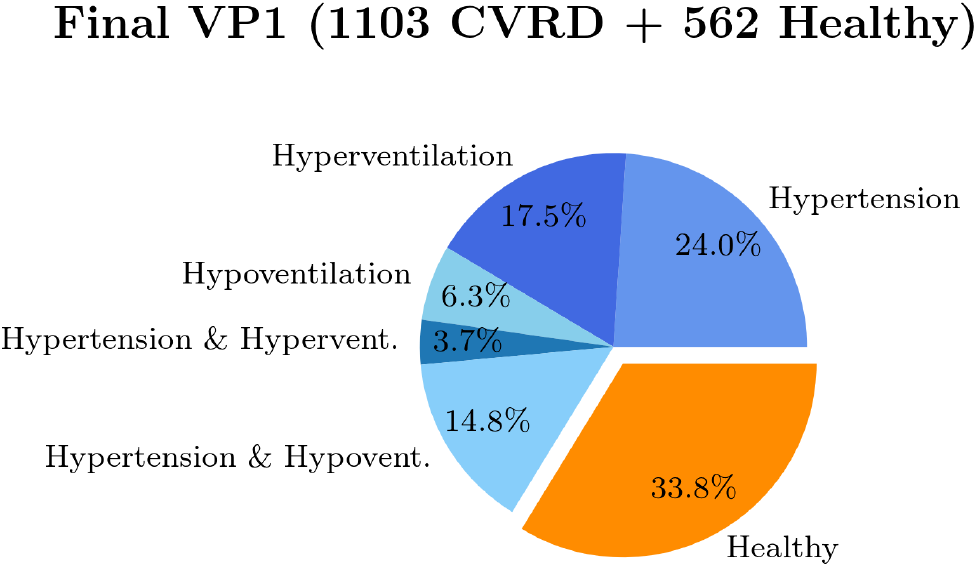
CVRD classification. CVRD classifications among the simulations that have not failed the filter criteria of VP 1.

Overall, the distribution ranges of the variables ensure a good level of heterogeneity, contributing to a diverse and realistic representation of the selected outputs. This allowed us to simulate a variety of scenarios, including a sufficient spectrum of pathological conditions.

Lastly, we report a quantitative comparison between the final VPs, 1 and 2, after applying the filter criteria. Of the 4096 simulations of VP 2, 1299 samples were accepted after applying the filtering criteria specified in Table 3. Table 7 compares the final VPs in terms of means, SDs, minimums, and maximums. We observe that the descriptive statistics are similar across all variables in both populations, with minor variations due to differences in the parameter selection used for their generation. Fig. 9 displays the Pearson correlation coefficients between the four wearable-derived signals and all the in-hospital variables for both final VPs. Table 8 shows the absolute sum of the wearable-derived Pearson correlation coefficients for all the in-hospital variables, for both VPs. For most in-hospital variables, the differences in absolute sums of correlations between the two VPs were small, with variations remaining below 0.09. However, Fig. 9 highlights stronger correlations between CSBP and CDBP with CVP in VP 1. Specifically, from Table 8 we can observe that the absolute sum of Pearson correlation coefficients for CVP in VP 1 is 1.90, compared to 1.57 in VP 2, reflecting an absolute difference of 0.33.

**Table 6.** Local sensitivity analysis for the cardio-respiratory model.

| Variable | Rank 1 | Rank 2 | Rank 3 | Rank 4 | Rank 5 |
| --- | --- | --- | --- | --- | --- |
| HR | $V_{\text{tot}}$ (-1.0) | $V_{\text{u,ven}}$ (0.5) | $G_{T_v}$ (-0.5) | $T_0$ (-0.4) | $hgb$ (-0.3) |
| CSBP | $V_{\text{tot}}$ (2.1) | $V_{\text{u,ven}}$ (-1.2) | $C_{\text{sat,O}_2}$ (-0.6) | $h_{\text{CO}_2}$ (-0.5) | $P_n$ (0.4) |
| CDBP | $V_{\text{tot}}$ (1.8) | $V_{\text{u,ven}}$ (-1.0) | $C_{\text{sat,O}_2}$ (-0.9) | $h_{\text{CO}_2}$ (-0.8) | $P_n$ (0.6) |
| $S_{\text{a,O}_2}$ | $hgb$ (-0.5) | $h_{\text{O}_2}$ (-0.1) | $F_{\text{iO}_2}$ (0.1) | $V_{\text{tot}}$ (-0.1) | $k_{\text{O}_2}$ (-0.1) |
| CVP | $V_{\text{tot}}$ (9.6) | $V_{\text{u,ven}}$ (-5.7) | $k_{\text{E,RV}}$ (1.3) | $V_{\text{u,art}}$ (-1.1) | $P_l$ (1.1) |
| SV | $V_{\text{tot}}$ (2.9) | $V_{\text{u,ven}}$ (-1.7) | $k_{\text{E,LV}}$ (-0.4) | $V_{\text{u,art}}$ (-0.3) | $k_{\text{E,RV}}$ (-0.3) |
| CO | $V_{\text{tot}}$ (2.1) | $V_{\text{u,ven}}$ (-1.2) | $k_{\text{E,LV}}$ (-0.3) | $T_0$ (-0.3) | $G_{T_v}$ (-0.3) |
| EF | $t_{\text{LA}}^c$ (1.5) | $T_0$ (-1.4) | $G_{T_v}$ (-1.4) | $P_{0,\text{LV}}$ (1.1) | $k_{\text{close,MV}}$ (1.1) |
| $P_{\text{a,O}_2}$ | $F_{\text{iO}_2}$ (1.1) | $C_{\text{sat,O}_2}$ (-0.9) | $V_{\text{tot}}$ (-0.9) | $P_{\text{atm}}$ (0.8) | $h_{\text{CO}_2}$ (-0.7) |
| $P_{\text{a,CO}_2}$ | $C_{\text{sat,O}_2}$ (-1.6) | $h_{\text{CO}_2}$ (-1.5) | $k_{\text{CO}_2}$ (0.9) | $\alpha_{\text{CO}_2}$ (0.7) | $\beta_{\text{CO}_2}$ (-0.6) |

**Table 7.** Comparison between the final VPs 1 and 2.

| Variable | Mean |  | SD |  | Min |  | Max |  | Unit |
| --- | --- | --- | --- | --- | --- | --- | --- | --- | --- |
|  | VP 1 | VP 2 | VP 1 | VP 2 | VP 1 | VP 2 | VP 1 | VP 2 |  |
| HR | 63.3 | 63.7 | 7.0 | 7.3 | 50.0 | 50.0 | 90.0 | 89.0 | beats · min<sup>-1</sup> |
| CSBP | 112.4 | 111.4 | 13.2 | 13.0 | 84.8 | 84.4 | 147.2 | 143.4 | mmHg |
| CDBP | 78.4 | 78.0 | 9.7 | 9.3 | 59.2 | 59.6 | 107.8 | 103.7 | mmHg |
| CPP | 34.0 | 33.3 | 5.4 | 5.5 | 17.6 | 17.4 | 42.7 | 42.6 | mmHg |
| MAP | 94.6 | 93.9 | 11.3 | 11.0 | 74.1 | 74.0 | 126.9 | 122.8 | mmHg |
| CVP | 3.2 | 3.3 | 1.5 | 1.8 | 0.0 | 0.0 | 7.3 | 8.8 | mmHg |
| SV | 103.3 | 101.3 | 17.1 | 17.4 | 52.2 | 51.4 | 128.6 | 128.6 | mL |
| CO | 108.5 | 106.7 | 14.5 | 14.7 | 72.2 | 73.1 | 145.7 | 146.3 | mL · s<sup>-1</sup> |
| EF | 62.5 | 62.2 | 2.5 | 2.7 | 51.5 | 51.6 | 67.8 | 67.6 | % |
| S<sub>a,O<sub>2</sub></sub> | 96.4 | 96.4 | 1.2 | 1.2 | 93.0 | 92.6 | 98.4 | 98.5 | mmHg |
| P<sub>a,O<sub>2</sub></sub> | 88.7 | 88.9 | 11.7 | 11.6 | 70.0 | 70.0 | 110.0 | 110.0 | mmHg |
| P<sub>a,CO<sub>2</sub></sub> | 40.0 | 40.6 | 5.1 | 5.3 | 30.0 | 30.0 | 48.1 | 49.5 | mmHg |

Means, SDs, minimums and maximums of variables of interest of final VPs 1 and 2.. Abbreviations: HR: heart rate; CSBP/CDBP: central systolic/diastolic blood pressure; CPP: central pulse pressure; MAP: mean arterial pressure; CVP: central venous pressure; SV: left ventricle stroke volume; CO: left ventricle cardiac output; EF: left ventricle ejection fraction; S<sub>a,O<sub>2</sub></sub>: arterial O<sub>2</sub> saturation; P<sub>a,O<sub>2</sub></sub>: arterial partial pressure of O<sub>2</sub>; P<sub>a,CO<sub>2</sub></sub>: arterial partial pressure of CO<sub>2</sub>.

**Table 8.** Wearable-derived and in-hospital variables correlations sums.

| Dataset | CVP | SV | CO | EF | P<sub>a,O<sub>2</sub></sub> | P<sub>a,CO<sub>2</sub></sub> |
| --- | --- | --- | --- | --- | --- | --- |
| VP 1 | 1.90 | 2.03 | 1.74 | 1.28 | 0.91 | 0.91 |
| VP 2 | 1.57 | 2.10 | 1.76 | 1.37 | 0.95 | 0.95 |

Absolute sums of the wearable-derived Pearson correlation coefficients for the in-hospital variables, for the final VPs 1 and 2. Abbreviations: CVP: central venous pressure; SV: left ventricle stroke volume; CO: left ventricle cardiac output; EF: left ventricle ejection fraction; P<sub>a,O<sub>2</sub></sub>: arterial partial pressure of O<sub>2</sub>; P<sub>a,CO<sub>2</sub></sub>: arterial partial pressure of CO<sub>2</sub>.

**Fig 9.**
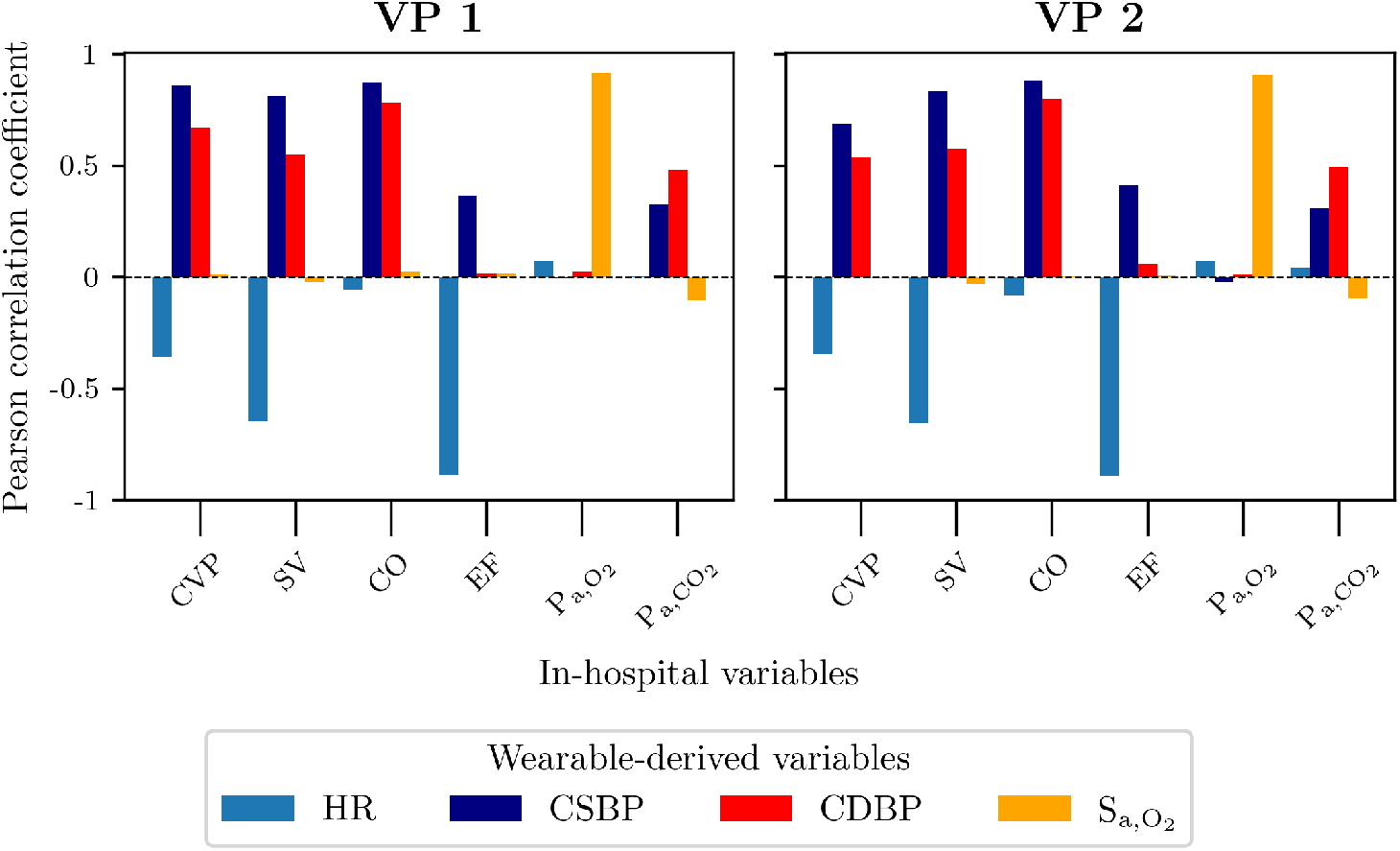
Wearable-derived and in-hospital variables correlations. Pearson correlation coefficients (y-axes) between the wearable-derived signals (HR, CSBP, CDBP, 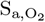) and the in-hospital ones (x-axes) for the final VPs 1 and 2. Abbreviations: HR: heart rate; CSBP/CDBP: central systolic/diastolic blood pressure; 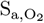: arterial O_2_ saturation; CVP: central venous pressure; SV: left ventricle stroke volume; CO: left ventricle cardiac output; EF: left ventricle ejection fraction; 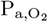: arterial partial pressure of O_2_; 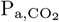: arterial partial pressure of CO_2_.

### GPR models predictions

Fig. 10 shows GPR model predictions versus true data for the in-hospital variables on the final VP 1 test set (333 samples). The model demonstrated strong predictive performance across all variables, particularly for cardiovascular variables such as CVP, SV, CO, and EF. In contrast, the predictions for the cardio-respiratory variables, i.e. 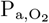 and 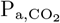, were less precise, as indicated by lower R^2^ values. These findings align with the results in Table 8, where the absolute sums of the Pearson correlation coefficients for the cardiovascular variables in final VP 1 were significantly higher than those for the cardio-respiratory variables.

**Fig 10.**
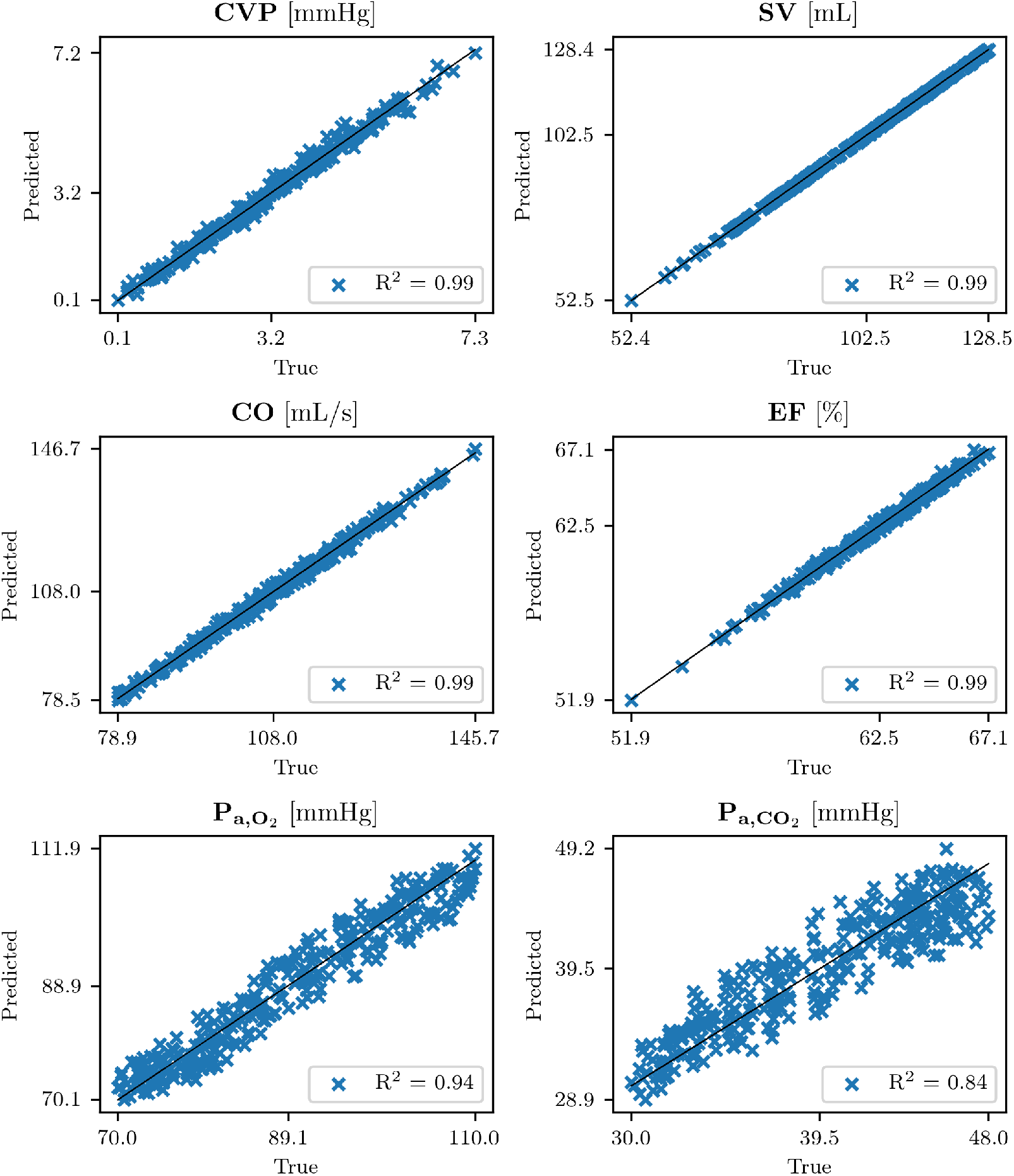
GPR models predictions versus true data. GPR models predictions (y-axes) versus true data (x-axes) for the final VP 1 test set, for all the in-hospital variables, together with the coefficient of determination R^2^. Abbreviations: CVP: central venous pressure; SV: left ventricle stroke volume; CO: left ventricle cardiac output; EF: left ventricle ejection fraction; 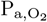: arterial partial pressure of O_2_; 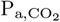: arterial partial pressure of CO_2_.

Table 9 summarizes the GPR model predictions for perturbed and non-perturbed input features across the final VP 1 test set (333 samples) and the final VP 2 dataset (1299 samples) for all the in-hospital variables. We recall that the training of the GPR models was done only on the training set of the final VP 1 (1332 samples).

**Table 9.** Performance of the GPR predictions.

| Target | Test set | Without perturbed inputs |  |  | With perturbed inputs |  |  |
| --- | --- | --- | --- | --- | --- | --- | --- |
| | | R<sup>2</sup> | maxRE | MRE | maxRE<sub>PI</sub> | MRE<sub>PI</sub> | CV (mean ± SD) |
| CVP | VP 1 | 0.99 | 86.1 % (0.5*) | 5.1 % (0.1*) | 81.1 % (0.6*) | 5.2 % (0.1*) | 48.2 ± 122.3 % |
| | VP 2 | 0.55 | 6761.5 % (4.2*) | 47.7 % (1.0*) | 7017.0 % (4.2*) | 48.0 % (1.0*) | 42.6 ± 216.7 % |
| SV | VP 1 | 0.99 | 0.6 % | 0.2 % | 0.6 % | 0.2 % | 8.5 ± 1.6 % |
| | VP 2 | 0.99 | 0.8 % | 0.2 % | 0.8 % | 0.2 % | 8.6 ± 1.7 % |
| CO | VP 1 | 0.99 | 2.9 % | 0.8 % | 2.6 % | 0.8 % | 8.9 ± 1.5 % |
| | VP 2 | 0.99 | 2.9 % | 0.7 % | 3.1 % | 0.8 % | 9.0 ± 1.6 % |
| EF | VP 1 | 0.99 | 1.0 % | 0.3 % | 0.9 % | 0.3 % | 2.1 ± 0.7 % |
| | VP 2 | 0.99 | 0.9 % | 0.2 % | 1.1 % | 0.2 % | 2.1 ± 0.7 % |
| P<sub>a,O<sub>2</sub></sub> | VP 1 | 0.94 | 6.8 % | 2.7 % | 10.0 % | 3.2 % | 12.9 ± 3.2 % |
| | VP 2 | 0.94 | 9.9 % | 2.6 % | 13.9 % | 3.3 % | 12.7 ± 3.3 % |
| P<sub>a,CO<sub>2</sub></sub> | VP 1 | 0.84 | 13.6 % | 4.2 % | 13.9 % | 4.4 % | 13.8 ± 3.5 % |
| | VP 2 | 0.91 | 13.1 % | 2.9 % | 16.2 % | 4.0 % | 13.5 ± 3.4 % |

Metrics and scoring of the GPR models predictions, with and without perturbed input data, on both the final VPs 1 and 2. Abbreviations: R<sup>2</sup>: coefficient of determination; maxRE: percentage maximum relative error of the GPR models predictions on test set; MRE: percentage mean relative error of the GPR models predictions on test set; maxRE<sub>PI</sub>: percentage maximum relative error of the stochastic distribution of GPR models predictions with perturbed inputs on test set; MRE<sub>PI</sub>: percentage mean relative error of the stochastic distribution of GPR models predictions with perturbed inputs on test set; CV: coefficient of variation of the stochastic distribution of GPR models predictions with perturbed inputs on test set; CVP: central venous pressure; SV: left ventricle stroke volume; CO: left ventricle cardiac output; EF: left ventricle ejection fraction; P<sub>a,O<sub>2</sub></sub>: arterial partial pressure of O<sub>2</sub>; P<sub>a,CO<sub>2</sub></sub>: arterial partial pressure of CO<sub>2</sub>. Unit: \* = mmHg.

For non-perturbed inputs, the table reports the following metrics: R^2^, maxRE (Eq. 7), and MRE (Eq. 8). For perturbed inputs, the table includes maxRE_PI_ (Eq. 13), MRE_PI_ (Eq. 14), as well as the means and SDs of **CV** (Eq. 15).

When comparing the GPR models performance between the final VPs 1 and 2, as reported in Table 9, negligible differences were observed for SV, CO, EF and 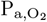. Slightly larger differences were noted for 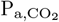, although the overall performance remains comparable. In contrast, the GPR model performance for CVP was significantly worse on the final VP 2. This aligns with the observation that the final VP 1 exhibited stronger correlations between the GPR input feature vector, i.e., the wearable-derived signals, and CVP, compared to the final VP 2 (see Fig. 9 and Table 8).

When comparing the performance of the GPR models with perturbed and non-perturbed input data, as reported in Table 9, we observed negligible differences in the results. This is further illustrated in Fig. 11, which shows the relative errors for the GPR models across all predicted variables in the final VP 1 test set. The figure compares the distributions of **RE** (relative errors for non-perturbed data, shown in blue) and **RE**_PI_ (relative errors for perturbed data, shown in orange). For each distribution, the means and SDs are displayed above the mean lines, while the maximum relative errors are indicated at the top. These plots demonstrate that the performance of the GPR models remains consistent between perturbed and non-perturbed input data, indicating that the method remains robust and performs well even when physiological perturbations are applied to the input data.

**Fig 11.**
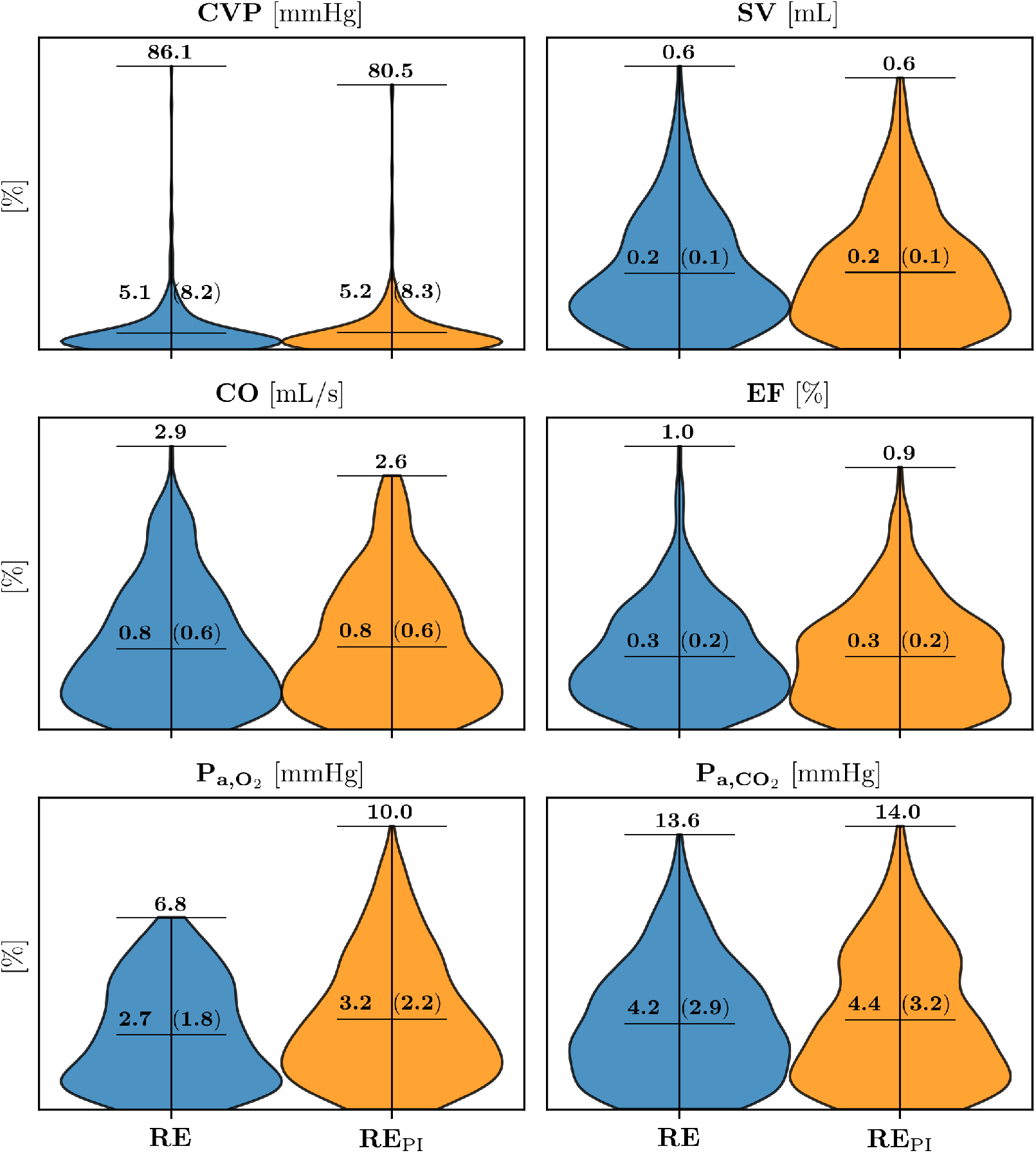
Relative errors of the GPR models predictions. Relative errors with and without perturbed inputs (orange and blue distributions respectively), for the final VP 1 test set. Over each distributions we annotate the mean and the SD, between round brackets, of the error, together with the maximum error, at the top of each distribution. Abbreviations: RE: percentage relative error of the GPR models predictions without perturbed inputs; RE_PI_: percentage relative error of the stochastic distribution of GPR models predictions with perturbed inputs; CVP: central venous pressure; SV: left ventricle stroke volume; CO: left ventricle cardiac output; EF: left ventricle ejection fraction; 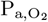: arterial partial pressure of O_2_; 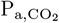: arterial partial pressure of CO_2_.

Fig. 12 shows the learning curves of maxRE for all GPR models trained with 100, 200, 300, 500, and 1000 samples drawn from the final VP 1 training set. The results indicate that the performance of the GPR models improves significantly with a training set size of at least 300 samples, suggesting that a minimum of 300 training samples is required to ensure reliable model performance.

**Fig 12.**
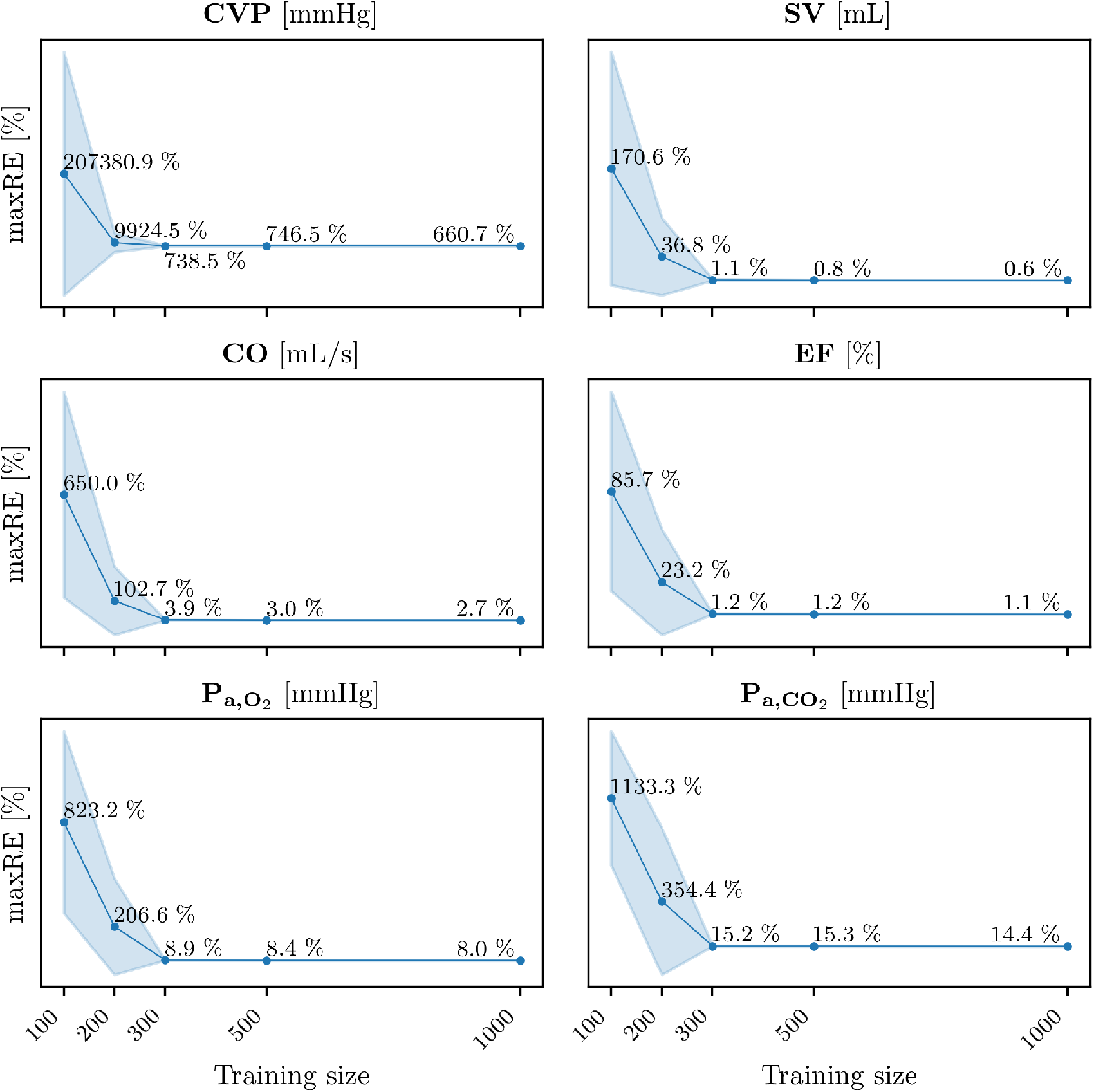
Learning curves of the GPR models. Learning curves of the percentage maximum relative errors of the GPR models predictions without perturbed input data (y-axes) for 100, 200, 300, 500 and 1000 training sizes sampled from the final VP 1 training set (x-axes). Abbreviations: maxRE: percentage maximum relative error of the GPR models predictions without perturbed inputs; CVP: central venous pressure; SV: left ventricle stroke volume; CO: left ventricle cardiac output; EF: left ventricle ejection fraction; 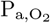: arterial partial pressure of O_2_; 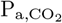: arterial partial pressure of CO_2_.

## Discussion

### Virtual database

In this study, we presented a methodology for generating a VP dataset that includes both healthy individuals and patients with CVRD, such as hypertensive patients, hyper- or hypo-ventilating ones, or combinations of those (see Fig. 8). This was achieved using an advanced computational cardio-respiratory model capable of simulating a wide range of clinically relevant variables. Although there are existing studies focused on generating virtual populations [24, 37, 38, 69, 70], to the best of our knowledge, our dataset is the most comprehensive in terms of the number of features it includes, spanning both cardiovascular and respiratory characteristics.

One of the key strengths of our approach is its speed and efficiency, in terms of time and computational cost. One simulation runs in 3 min and 45 s on a system equipped with an AMD^®^ Ryzen™ Threadripper™ 3990X processor, featuring 64 cores and 128 threads, which allowed us to generate up to 5000 virtual patients in approximately 15 hours, with 20 simulations running concurrently. This ability to quickly generate vast datasets opens up new opportunities for predictive modeling, particularly when real patient data may not be available in sufficient quantity or quality.

Clinical datasets often predominantly consist of patients who are hospitalized, typically under the effect of drugs, or undergoing surgery, thus in advanced stages of disease [11–14, 33, 34]. This makes it challenging to study predictive models for a healthy population or individuals with less acute conditions, such as those with early-stage forms of disease. In contrast, our dataset specifically describes these conditions, including healthy states and mild to moderate CVRD.

Moreover, our VP generation methodology could be used in the future to simulate a broader variety of CVRD, expanding the selection and refinement of parameter ranges and their combinations. This could also include wave propagation models for the circulatory system, such as those in [71, 72], which would allow addressing additional pathological aspects and enable more targeted predictive studies, as in [37, 69, 70]. This would not only allow for the inclusion of additional conditions but also enable the modeling of different levels of severity within specific diseases, thus providing a more nuanced dataset for predictive studies. By designing and simulating more targeted scenarios, this approach could enhance the development of more accurate, robust, and clinically relevant predictive models.

### Prediction of in-hospital variables

This study demonstrates the potential of using virtual patient data to predict in-hospital variables, such as CVP, CO, SV, EF, 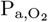 and 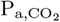, from signals that are acquirable through wearable devices, i.e. HR, BPs and 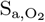. In this in silico context, GPR emerges as a particularly suitable predictive methodology. Importantly, these predictions show good accuracy, even when accounting for potential errors introduced by wearable signal acquisition, as shown in Fig. 11. Specifically (see Table 9), the mean relative errors for non-perturbed data are as follows: CVP: 5.1 % (0.1 mmHg), SV: 0.2 %, CO: 0.8 %, EF: 0.3 %, 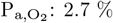, and 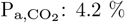. When comparing these results to those obtained with perturbed inputs, the differences were negligible: CVP: 5.2 % (0.1 mmHg), SV: 0.2 %, CO: 0.8 %, EF: 0.3 %, 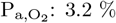, and 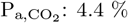. Cardiovascular parameters like CVP, SV, CO, and EF, show consistently high accuracy. Whereas, the predictions for 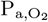 and 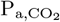 were slightly less precise but still demonstrate good accuracy.

In terms of clinical acceptability, there is no universal definition of error thresholds, as they vary depending on the variable being measured and the methodology in use. However, considering that wearable devices primarily serve as screening tools rather than diagnostic instruments, a certain degree of error is generally tolerated. Within this context, the error thresholds observed in our study can be considered clinically acceptable, as they align with the intended purpose of these devices in preliminary assessment and continuous monitoring.

Moreover, to ensure that the predictive capabilities of our model are not overly dependent on the specific parameter choices that we made for the generation of the VP, we tested the GPR models, which were trained on the VP 1 dataset, on a new dataset, VP 2, obtained varying different combinations of parameters. In particular, the mean relative error differences between the performance on VP 1 and VP 2, for the variables predicted with perturbed input, are (see Table 9): SV (0.0 %), CO (0.1 %), EF (0.1 %), 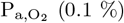, followed by 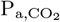, and finally CVP (0.9 mmHg). For all the predicted variables, with the exception of CVP, the mean relative error differences are negligible. These results suggest that the model’s predictive performance is not overly reliant on the specific characteristics of the dataset used for training, which provides confidence that the model can be effectively applied to different, potentially real-world, datasets.

One of the critical factors enabling the high accuracy of predictions in this study is the large dataset of virtual patients generated through our computational framework. Results reported in Section GPR models predictions revealed that at least 300 training samples are required to achieve satisfactory predictions. However, generating such a large dataset using real-world signals is often time-consuming, resource-intensive, or even infeasible due to logistical and ethical constraints. To address this limitation, future research could explore the combination of real-world data with in silico generated data for training GPR models, as explored in [73, 74]. By augmenting real patient datasets with virtual data, it might be possible to develop predictive models that perform well on real-world data while reducing the dependence on extensive real datasets. This hybrid approach could facilitate the creation of accurate and clinically relevant predictive tools, particularly in scenarios where data collection is challenging or constrained.

## Limitations

The results presented in this study were obtained entirely in an in silico setting. Although the computational framework and generated datasets have been rigorously tested, real-world validation is necessary to confirm the clinical utility and reliability of the predictive models.

In the local sensitivity analysis performed on the cardio-respiratory model, each parameter is varied by 20 % to assess its impact on the clinically relevant variables. However, this approach does not account for the fact that some parameters may exhibit greater or smaller physiological variability. Assigning physiologically accurate ranges to each model parameter is challenging, particularly considering the large number of parameters involved (286). Additionally, the local sensitivity analysis does not capture potential interactions between parameters, which could also influence the model’s outputs. Therefore, while the local sensitivity analysis provides a computationally efficient way to assess individual parameter impacts, a more comprehensive approach would involve performing a global sensitivity analysis, and a more refined selection of parameter variability ranges. This would ultimately lead to a more accurate choice of parameters for the generation of the VP. However, such approach would demand significantly more computational resources and an optimized methodology to remain feasible.

The virtual dataset is generated by varying the most significant model parameters within physiologically plausible ranges and, after applying the filter criteria, all virtual variables align with physiological or pathological ranges. However, the cardio-respiratory model has not been specifically parameterized to reproduce full pathological profiles. Instead, virtual subjects were identified as pathological based solely on certain output variables exceeding literature-reported threshold values. Consequently, we have not verified whether their overall physiological profile was consistent with that of a real patient.

## Conclusion

This study highlights the potential for predictive studies on remote and continuous monitoring of both healthy individuals and those affected by cardiovascular and respiratory diseases, using virtual patient datasets generated through an advanced computational cardio-respiratory model. The ability to simulate a broad range of physiological conditions, provides a resource for developing and testing predictive tools, especially in scenarios where real-world data is limited or unavailable. A critical advantage of this methodology is its ability to generate large, diverse datasets in a computationally efficient manner, facilitating the development of predictive models that require substantial training data.

The results demonstrate the high accuracy of GPR-based predictions for key cardiovascular and respiratory parameters, even under conditions of perturbed input data. Moreover, the negligible differences observed when testing models trained on one virtual patient dataset against a second independently generated dataset, indicate that the predictive capabilities are not overly dependent on the specific parameter choices used to generate the virtual population. This suggests that the proposed approach is generalizable across different datasets.

Future efforts should focus on refining the parameter ranges and combinations to expand the variety of simulated conditions, as well as on testing the methodology on real patient data, exploring also the possibility of an hybrid approach with both real and in silico data.

## Supporting information

**S1 Appendix. Cardio-respiratory model equations and validation**. The model is a non-linear system of differential-algebraic equations describing cardiovascular functions, including blood circulation, respiration, gas transport, and short-term regulation.

**S2 Appendix. Definition of cardiovascular and cardio-respiratory indexes**. A precise description of the computation of the extracted model output variables.

**S3 Appendix. Virtual population 2 results**. Convergence, distributions, post-processing filter and CVRD-classification.

## Data Availability Statement

The cardio-respiratory model code is publicly available at https://doi.org/10.5281/zenodo.14975200. Simulation input files and generated data of the virtual population are available from https://doi.org/10.5281/zenodo.14962119.

## Author contributions

**Conceptualization:** Bianca Maria Laudenzi, Lucas Omar Müller

**Data Curation:** Bianca Maria Laudenzi, Lucas Omar Müller

**Formal analysis:** Bianca Maria Laudenzi, Lucas Omar Müller

**Funding acquisition:** Lucas Omar Müller.

**Investigation:** Bianca Maria Laudenzi

**Methodology:** Bianca Maria Laudenzi, Lucas Omar Müller, Alberto Cucino, Sergio Lassola, Eleonora Balzani

**Project administration:** Lucas Omar Müller

**Resources:** Lucas Omar Müller

**Software:** Bianca Maria Laudenzi

**Supervision:** Lucas Omar Müller

**Validation:** Bianca Maria Laudenzi, Lucas Omar Müller, Alberto Cucino, Sergio Lassola, Eleonora Balzani

**Visualization:** Bianca Maria Laudenzi

**Writing – original draft**: Bianca Maria Laudenzi

**Writing – review & editing**: Lucas Omar Müller, Alberto Cucino, Sergio Lassola, Eleonora Balzani

